# Identification of Myocardial Infarction (MI) Probability from Imbalanced Medical Survey Data: An Artificial Neural Network (ANN) with Explainable AI (XAI) Insights

**DOI:** 10.1101/2024.02.28.24303497

**Authors:** Simon Bin Akter, Sumya Akter, Tanmoy Sarkar Pias, David Eisenberg, Jorge Fresneda Fernandez

## Abstract

In the healthcare industry, many artificial intelligence (AI) models have attempted to overcome bias from class imbalances while also maintaining high results. Firstly, when utilizing a large number of unbalanced samples, current AI models and related research have failed to balance specificity and sensitivity – a problem that can undermine the reliability of medical research. Secondly, no reliable method for obtaining detailed interpretability has been put forth when addressing large numbers of input features. The present research addresses these two critical research gaps with a proposed lightweight Artificial Neural Network (ANN) model. Using 43 input features from the 2021 Behavioral Risk Factor Surveillance System (BRFSS) dataset, the proposed model outperforms prior models in producing balanced outcomes from markedly unbalanced large survey data. The efficacy of this proposed ANN model is attributed to its simplified design, which reduces processing demands, and its resilience in identifying the probability of myocardial infarction (MI). This is demonstrated by its 80% specificity and 77% sensitivity, and is substantiated by a Receiver Operating Characteristic Area Under the Curve (AUC) of 0.87. The outcomes across the scopes of each specified data domain were also separately represented, thus demonstrating the proposed model’s robust sensitivity. The interpretability of the model, as measured by Shapley values, reveals substantial correlations between myocardial infarction (MI) and its risk factors, including long-term medical conditions, socio-demographic factors, personal health habits, economic and social status, healthcare availability and affordability, as well as impairment statuses, providing valuable insights for improved cardiovascular risk assessment and personalized healthcare strategies.

## Introduction

Cardiovascular diseases have emerged as a key global health concern, accounting for a considerable number of fatalities globally. Heart diseases, such as coronary artery disease, myocardial infarction, and congestive heart failure, are the most common and life-threatening of these disorders. The frequency of these diseases highlights the critical need for accurate and prompt diagnosis in order to enhance patient treatment and public health outcomes^1,2,3,4,5,6,7,8,9,10^. Medical surveys have been an important source of data for cardiovascular research, allowing for the gathering of a wide range of health indicators, lifestyle habits, genetic variables, and patients’ past medical records^11,12^. The study of such large datasets provides a chance to uncover critical risk variables, comprehend illness patterns, and create effective diagnostic strategies^13,14^. However, because of the enormous number and complexity of these datasets, advanced computational approaches are required to extract relevant insights.

Machine learning and deep learning breakthroughs in recent years have revolutionized the area of medical data analysis^15,16,17^. These strategies have shown great potential in a variety of healthcare applications, including medical picture analysis, disease diagnosis, drug development, and personalized medical care recommendations^18,19,20^. Support Vector Machines, Random Forests, Decision Trees, K-Nearest Neighbors, Gaussian Naïve Bayes, Gradient Boosting, Adaptive Boosting, Random Undersampling Boosting, and Logistic Regression are examples of machine learning algorithms that have proved successful in discovering patterns and predicting outcomes in medical datasets^21,22,23^. Deep learning models, specifically Convolutional Neural Networks (CNNs) and Recurrent Neural Networks (RNNs), have excelled in image identification, natural language processing, and sequential data analysis in the medical field^24,25^.

Much notable research has obtained high accuracy, albeit without additional analysis to ensure reliability. Sensitivity and specificity analysis are therefore important analyses that can help demonstrate such reliability and ensure balanced results. Of the prior research that did attempt such additional analyses, the sensitivity and specificity were ultimately imbalanced, thus leading to imbalanced results - a critical problem that the proposed model of the current paper resolves. For example, in an analysis by Akkaya et al.^26^ on heart disease prediction, Extreme Gradient Boosting (XGB) obtained the highest accuracy which was 0.90. However, the obtained sensitivity was 0.27, and the specificity was 0.94, showing this lack of balance^26^. In an analysis by Mamun et al.^27^ on heart disease prediction, Logistic Regression (LR) obtained its highest accuracy of 0.9157. However, they obtained an imbalanced sensitivity and specificity of 0.9232 and 0.5261, respectively^27^. Accordingly, in an analysis by Bhola et al.^28^ on chronic disease prognosis, Random Forest (RF) obtained their highest accuracy for heart disease of 0.9118^28^, yet neither sensitivity nor specificity analysis was performed. Likewise, Raghupathi et al.^29^ performed a study on preventive healthcare that analyzed the relationship between behavioral habits and chronic diseases, including variables from health behavior, chronic disease, and demographic section. In their analysis, the highest accuracy was obtained for heart disease, through the SPSS modeler and auto-classifier model, which was 95.1. Though not implemented by their study, their admirable accuracy alone does not demonstrate the overall performance of their models, since when accuracy is high, sensitivity and specificity could still be varied^26,27^. Moreover, such additional analyses could improve their study’s significance and reveal additional relationships between different risk factors. Hence, prior research has predominantly resulted in lower sensitivity or unbalanced outcomes for the target classes due to the unbalanced dataset^26,27^, often with no sensitivity and specificity even performed^28,29^.

For interpreting the reliability of artificial intelligence (AI) models, sensitivity can be considered the most important factor. Research by Nasimov et al. (2022) explored the use of feature importance when predicting chronic heart diseases with weighted K-Nearest Neighbors (KNN), obtaining the highest accuracy of 0.743. Their feature importance technique^30^ aimed to reduce variation in the results across different approaches, but failed to provide detailed information. Instead, their analysis only provided an overview of feature significance without revealing the significance based on the target classes separately. For this type of analysis, identifying the relevant features that influence the prediction of the particular target classes is essential for efficient model interpretation and decision-making. A statistical analysis by Park et al.^31^ revealed being underweight as an important risk factor for cardiovascular disease. However, artificial intelligence (AI) techniques could actually provide a more detailed clarification, with explainable AI (XAI) offering more detailed information about exactly why being underweight influenced cardiovascular disease for that dataset - an essential extra analysis that the current paper does perform.

Despite substantial advances in the field of cardiac disease identification using machine learning and deep learning, there are still difficulties to overcome. These difficulties highlight the necessity of this research in contributing to the current body of knowledge and proposing reliable methods to address the challenges identified in the literature review. Missing values and class imbalance are common issues in medical survey data, which can lead to biased conclusions and reduce the reliability of classifiers^26,27,28^. The interpretability and explainability of artificial intelligence (AI) models are critical in the medical arena since healthcare practitioners demand insights into the model’s decision-making process^32,33^. Deep learning models, particularly neural networks, have been criticized for their “black-box” nature, which makes it difficult to grasp the logic behind the predictions made by these approaches^34,35,36,37,38,39,40^. This study intends to overcome these important issues by proposing reliable, explainable, and thus more transparent methods for exploring cutting-edge deep-learning techniques for medical research and practice. These techniques expand the current body of knowledge, increase the specificity, sensitivity, and interpretability of the models, and ultimately can improve the quality of healthcare delivery and cardiovascular clinical practice by improving myocardial infarction (MI) probability identification using medical survey data.

The present research looks at the way machine learning and deep learning techniques can identify the probability of myocardial infarction (MI) using medical survey data. The primary goal is to create a dependable and automated model that will help healthcare providers properly diagnose myocardial infarction (MI) probability at an early stage. Identification of essential risk factors and biomarkers associated with heart disease has been performed by leveraging information from medical surveys. A very large in-depth survey instrument collected in 2021 that included a range of subjects, including medical history, lifestyle, diet, and health practices was analyzed in this study. 43 pertinent characteristics were chosen among over 300 parameters based on the previous studies, with the main goal of predicting myocardial infarction (MI) probability. The rigorous preparation of the data included cleaning missing data and outliers. The outcomes of multiple machine learning and deep learning models were compared. A reliable lightweight architecture of Artificial Neural Networks (ANNs) was proposed to identify the probability of myocardial infarction (MI). Despite the erroneous complexity of the data, the model was still able to yield very reliable outcomes. Interpretability^41^ and sensitivity^42^ analysis approaches were employed to ensure that the model’s predictions were explainable and therefore most practically useful in medical practice^43^, thus showing the distinct contributions of each input domain to the predictive framework.

The key contributions and identified observations are summarized below.

- This is the first time that an exploration of the myocardial infarction (MI) probability prediction with interpretability and sensitivity was implemented using diverse data domains.
- An ANN model has been proposed that takes less computational time due to its light architecture and it can properly handle unbalanced outcomes [specificity: 80% and sensitivity: 77%] from heavily imbalanced survey data (tested on: 107829 healthy cases and 4964 MI cases).
- The interpretability of the proposed artificial neural network (ANN) model, as measured by Shapley values^44^, reveals noteworthy relationships. As an example, individuals with heart disease and stroke history have a comparative probability above 0.40 and 0.25 respectively of having a myocardial infarction (MI).

In summary, this study addresses the essential issue of myocardial infarction (MI) identification by using deep learning algorithms from medical survey data. This study hopes to improve healthcare outcomes by contributing to the development of strong diagnostic approaches, opening the path for early identification and successful management of cardiac disorders. The next section addresses the most relevant literature on healthcare improvement using patients’ records. The literature review section describes the recent relevant works on chronic disease predictions from patients’ records incorporating the input features impact and also mentions the limitations where presence of scope for improvement. Following, the dataset section provides information on the total number of samples, distribution of target classes, and rationale for selecting this dataset version. Consecutively, the methodology section describes the complete technical workflow of the proposed approach, where each sub-approach has been mentioned to obtain the final outcomes. Later on, feature selection provides the list of relevant features that have been selected based on past studies and also provides statistical measures like the Chi-squared range and P-value range to show the relevance of the specified input features with the target. The dataset contains 18 times more healthy cases (413207 records) than MI cases (22831 records), resulting in an imbalance. Without addressing the class imbalanced, the initial highest sensitivity (0.67) was significantly lower than the specificity (0.98) due to the nature of the data. Sequentially, the data preprocessing section describes the techniques such as outliers and missing values handling, label encoding, and Minority-weighted Sampling to prepare this large dataset to properly train the artificial intelligence models for predicting the target with the highest outcomes. After cleaning outliers and missing values, the dataset size was reduced by 52.62% (from 438693 records to 230845 records). The dataset that has been used in this study has multiple input domains that provide patients’ information on socio-demographic and economic factors, medical and impairment conditions, healthcare accesses, health habits, and preventive health services. The sensitivity analysis provides outcomes with different performance matrices on how these feature groups individually perform to predict the target. Following, the interpretability analysis section shows the impact of each input feature to predict the target class using SHapley Additive exPlanations (SHAP) values. SHAP helps to understand how a model made a specific prediction by measuring its impact on predictions in different combinations, based on cooperative principles. The sensitivity and interpretability analysis contributed to identifying the risk factors for myocardial infarction (MI) by quantifying the impact of the features. Subsequently, the experiment and result analysis section describes the obtained results using different performance matrices for all the applied approaches. In this way, the performance of the proposed approach can be compared with the existing approaches. The final result has a balanced specificity of 0.80 and a sensitivity of 0.77 by addressing class imbalance. Finally, the discussion section summarizes all the findings and observations derived from the study in a comprehensive manner.

## Literature review

The integration of Artificial Intelligence (AI) with comprehensive data holds enormous potential for improving cardiac disease prediction and preventative healthcare methods. These prediction models could utilize massive volumes of data to deliver personalized risk assessments, improve real-time monitoring of heart disease risk factors, and enable large-scale screening to identify high-risk groups quickly by using the capability of AI algorithms^45,46,47^. Regardless of the potential benefits, ethical concerns, data quality issues, and validation issues must be carefully addressed to guarantee the responsible and fair use of AI technology in healthcare. The effective deployment of data integration through AI has the potential to transform healthcare practices from reactive to proactive approaches, resulting in earlier identification, lower healthcare expenditures, and improved patient outcomes globally.

A predictive model for heart disease identification based on a two-level stacking of several classifiers (base level and meta-level) was proposed by Mohapatra et al.^48^. The UCI dataset was used to discover significant data trends, allowing machine learning algorithms to enhance predictions in the healthcare sector. The obtained accuracy, precision, sensitivity, and specificity for the proposed approach were respectively 0.92, 0.926, 0.926, and 0.909. This approach was tested on 33 healthy and 41 heart disease cases. The study conducted by Mohan et al.^49^ proposed an innovative way of predicting cardiovascular disease using machine learning techniques using the UCI dataset. The strategy focuses on finding relevant traits in order to increase prediction accuracy. The prediction model includes many characteristics and classification methodologies, resulting in an improved performance level. The obtained accuracy, precision, sensitivity, and specificity for the proposed approach were respectively 0.884, 0.901, 0.928, and 0.826. The analysis included multi-class prediction from class 0 to class 4 indicating no heart disease risk to higher risk of heart disease risk. However, the proposed study was conducted using 303 records. To predict cardiac disease, the study by Asif et al.^50^ proposed an ensemble learning (extra tree classifier, random forest, XGBoost, and CatBoost) model that integrates multiple preprocessing stages, hyperparameter optimization approaches, and ensemble learning algorithms. Multiple datasets from Kaggle have been merged to conduct this analysis. The obtained accuracy, precision, and specificity for the proposed approach were respectively 0.9815, 0.9508, and 0.9809. Data from three datasets including the first dataset: 297 records, the second dataset: 1025 records, and the third dataset: 303 records were used in this study. Akkaya et al.^26^ analyzed survey data from the BRFSS 2020 using eight different classification algorithms, including LR, SVM, NB, K-NN, DT, Adaboost, MLP, and XGB to detect cardiac diseases at an early stage. This study employed the SMOTE-Tomek Link approach to generate synthetic data and balance the dataset since the data exhibited an unbalanced distribution of the heart disease variable. Outlier analysis was conducted to classify the data as outliers or non-outliers. After preprocessing, 280293 records (tested on: 51884 healthy and 4175 heart disease cases) were used for this study. The obtained accuracy, sensitivity, and specificity from the proposed approach were 0.9, 0.27, and 0.94, respectively. Mamun et al.^27^ analyzed survey data from the BRFSS 2020 using six different machine learning algorithms, including Xgboost, Adaboost, Random Forest, Decision Tree, Logistic Regression, and Naïve Bayes to predict heart disease at an early stage. In this study, 319795 records were used. The obtained accuracy, sensitivity, and specificity from the proposed approach were 0.9157, 0.9232, and 0.5261, respectively.

Bhola et al.^28^ used machine learning algorithms to predict five main chronic diseases including heart disease, arthritis, pulmonary disease, renal disease, and diabetes based on behavioral risk variables using a single dataset from the BRFSS database in 2017. The obtained accuracy for heart disease prediction using the proposed approach was 0.9118. A chatbot was proposed based on the study to identify disease incidence and give personalized advice to reduce risk through interactive data visualization. The study conducted by Raghupathi et al.^29^ looked at the relationship between behavioral habits and chronic illnesses using data from the Behavioural Risk Factor Surveillance System (BRFSS) database for 2012. The study reveals substantial positive and negative connections between specific chronic illnesses and behavioral patterns using neural networks in SPSS Modeller. The highest accuracy from the proposed approach was 0.951. The research study conducted by Jindal et al.^51^ focused on applying machine learning methods such as Logistic Regression and K-Nearest Neighbour to predict cardiac illnesses based on various medical parameters using the UCI dataset. When compared to earlier classifiers such as Naive Bayes, the proposed heart disease prediction approach performed considerably. The highest accuracy for heart disease prediction using the proposed approach was 0.885. The study implemented by Singh et al.^52^ highlights the vital role of the heart in living creatures as well as the rising number of heart-related disorders that cause weariness or death using the UCI dataset. The research presents a prediction system based on machine learning methods such as K-Nearest Neighbour, Decision Tree, Linear Regression, and Support Vector Machine to address this issue. The highest accuracy from the proposed approach was 0.83. Using the UCI heart disease dataset, Kavitha et al.^53^ proposed a unique machine-learning technique for predicting cardiovascular disease. Their research included regression and classification approaches, including three machine learning algorithms: Random Forest, Decision Tree, and a Hybrid model (RF and DT). The highest accuracy from the proposed approach was 0.887. The study conducted by Shah et al.^54^ looks at the way data mining and machine learning approaches may be used to predict cardiac disease. The researchers examined four supervised learning algorithms on a dataset of 303 heart disease patients from the UCI database including Naïve Bayes, Decision Tree, K-Nearest Neighbour, and Random Forest. The highest accuracy from the proposed approach was 0.9079. The study by Repaka et al.^55^ offers Smart Heart Disease Prediction (SHDP), a web tool that uses data mining techniques (AES-encrypted data transport), and notably Navies Bayesian classification. Based on standardized medical profile data (collected), the algorithm predicted heart disease risk variables such as age, blood pressure, cholesterol, gender, and blood sugar. The highest accuracy from the proposed approach was 0.8977.

The study by Nasimov et al.^30^ provides an innovative way of identifying the value of numerous elements in predicting the influence on a patient’s health, particularly in situations of chronic disorders using Artificial Intelligence (AI) approaches. This analysis was conducted using the BRFSS dataset. The obtained accuracy from the proposed approach was 0.743. Existing approaches for measuring feature importance (FI) provide varied results, making interpretation problematic. The suggested strategy attempts to reduce the differences and produce more consistent results. The cross-sectional study provided by Park et al.^31^ analyzed data from the BRFSS database for over 491,000 US adults in 2013 to determine if underweight persons (BMI 18.5 kg/m2) had an independent risk for cardiovascular disease (CVD) using statistical analysis and interpretation approaches. According to the findings, the underweight group had a 19.7% greater risk of CVD than the normal-weight population.

The combination of Artificial Intelligence (AI) with comprehensive data constitutes a watershed moment in the field of cardiovascular disease prediction, with profound implications for preventative healthcare practices. The successful application of AI-survey data integration has the potential to catalyze a paradigm change in preventive healthcare practices, increasing early detection rates, optimizing resource allocation, and improving patient outcomes on a worldwide scale. A study conducted by Mohapatra et al.^48^ where precision, sensitivity, and specificity were reported as 0.926, 0.926, and 0.909, respectively. However, data for each predictive class was very limited and balanced (healthy: 33, heart disease: 41) in this study^48^. Similarly, Mohan et al.^49^ conducted a study where precision, sensitivity, and specificity were reported as 0.901, 0.928, and 0.826, respectively for five predicting classes. However, data used for this study^49^ was very limited (only 303 records were used in this study). Following, a study conducted by Asif et al.^50^ precision and specificity were reported as 95.08 and 98.09, respectively. However, the analysis^50^ was tested on very limited data (only 1625 records were used in this study). In real-world situations, there are frequently fewer cases of cardiac disease than healthy ones, resulting in an imbalance in the data. While a model may perform well in balanced circumstances, it might struggle in imbalanced situations in real life^26,27^. To truly assess the effectiveness of proposed approaches^48,49,50^, it is crucial to test them in unbalanced scenarios using a larger number of samples. The performance measurements across various unbalanced circumstances should be thoroughly described in order to appropriately assess the success of the suggested approaches in real-life situations. A study conducted by Akkaya et al.^26^ employed a large dataset (tested on: 51884 healthy and 4175 heart disease cases) where accuracy was reported as 0.89. However, the sensitivity (0.27) was significantly lower than the specificity (0.94) for this approach^26^. Similarly, Mamun et al.^27^ conducted a study using a large dataset (319795 records) where accuracy was reported as 0.9157. However, the specificity (0.5261) and sensitivity (0.9232) were unbalanced for this approach^27^. Most studies^28,29,30,51,52,53,54,55^ lack analysis regarding sensitivity and specificity, which are considered as crucial performance indicators for healthcare-related predictions. The studies conducted by Nasimov et al.^30^ and Park et al.^31^ analyzed the importance of input features in identifying risk factors, but the analyses lacked sufficient detail including sensitivity and interpretability analysis. To improve comprehension of these findings, more explanation and in-depth analysis are required.

The present study of the literature review on the application of artificial intelligence (AI) in medical diagnostics has produced useful insights, yet it appears to be lacking these mentioned important aspects that are required to implement for improving the application of AI in the cardiovascular domain. The prediction of myocardial infarction (MI) probability from medical survey records with diverse data domains related to the prediction target has not been explored yet in the present studies. To improve the reliability and applicability of such models few aspects are necessary to address which are also not thoroughly explored in the past literature. To begin, there is a notable lack of discussion of the obtained specificity and sensitivity of the proposed models in terms of predicting cardiovascular diseases using a larger number of samples under unbalanced circumstances. Secondly, there is a lack of comprehensive discussion of the interpretability and explicability of AI models in this study. Given its “black-box” nature, AI models are difficult to employ in clinical contexts. Thus, transparent model structures and explainable AI (XAI) approaches must be implemented^56^. Last but not least, sensitivity analysis to determine which data domains contribute the most to model predictions is conspicuously missing in this study. Addressing these flaws will not only improve the scientific study but will also ensure that artificial intelligence (AI) is used responsibly and effectively in medical diagnostics. The complete summary of the literature review is provided in **Table 1**.

**Table 1.** Overview of the literature review. The comprehensive summary of the literature review provided insights and synthesized information on the issue, laying the foundation for this analysis.

| Researcher | Year | Research Topic | Dataset | Modeling Tech-nique | Accuracy |
| --- | --- | --- | --- | --- | --- |
| Mohapatra et al.<sup>48</sup> | 2023 | Heart disease identi-fication | UCI | Two-level stacking | 92.00% |
| Asif et al.<sup>50</sup> | 2023 | Heart disease identi-fication | Kaggle datasets | EL (ETC, RF, XG-Boost, CatBoost) | 98.15% |
| Akkaya et al.<sup>26</sup> | 2022 | Heart disease identi-fication | BRFSS | XGB | 89.00% |
| Mamun et al.<sup>27</sup> | 2022 | Heart disease identi-fication | BRFSS | LR | 91.57% |
| Nasimov et al.<sup>30</sup> | 2022 | Chronic disorders identification | BRFSS | K-NN (weighted) | 74.30% |
| Bhola et al.<sup>28</sup> | 2021 | Chronic disease identification | BRFSS | LR, RF | 91.20% |
| Jindal et al.<sup>51</sup> | 2021 | Heart disease identi-fication | UCI | LR, K-NN | 88.50% |
| Kavitha et al.<sup>53</sup> | 2021 | Heart disease predic-tion | UCI | Hybrid model (LR, K-NN) | 88.70% |
| Shah et al.<sup>54</sup> | 2021 | Heart disease predic-tion | UCI | K-NN | 90.79% |
| Singh et al.<sup>52</sup> | 2020 | Heart-related disorders prediction | UCI | K-NN | 87.00% |
| Mohan et al.<sup>49</sup> | 2019 | Heart disease predic-tion | UCI | HRFLM | 88.70% |
| Repaka et al.<sup>55</sup> | 2019 | Heart disease predic-tion | Collected | GNB | 89.77% |
| Park et al.<sup>31</sup> | 2017 | Risk analysis for car-diovascular disease | BRFSS | Statistical analysis | Not applicable |
| Raghupathi et al.<sup>29</sup> | 2017 | Chronic illnesses prediction | BRFSS | Neural networks (SPSS) | 96.00% |

## Dataset

The present study dives into the early detection of myocardial infarction (MI) by an in-depth examination of 438693 individuals data from the survey conducted by the Centers for Disease Control and Prevention (CDC) on the Behavioral Risk Factor Surveillance System (BRFSS) from the year 2021^57^. This particular version of the BRFSS dataset has been chosen due to it has the most relevant features and the largest amount of data compared to other versions. The cardiovascular disease segment of the survey highlighted the key feature column that is figured to be of the utmost significance for myocardial infarction (MI) probability prediction, which has served as the basis for the target column. A thorough examination of the data demonstrates that the data distributions of class 0 (healthy cases) and class 1 (MI cases) are respectively around 94% and 5.21%, showing a persistent preponderance of class 0, which corresponds to outcomes associated with healthy individuals. The probability prediction model for myocardial infarction (MI) has been developed by examining the series worth of data. The research seeks to uncover critical risk factors and trends that could contribute to the prompt detection and prevention of myocardial infarction (MI), leading to the progress of public health initiatives and interventions.

## Methodology

Developing a prediction model to estimate the risk of myocardial infarction (MI) using extensive medical survey data and influential machine learning and deep learning techniques is crucial. The model’s astounding performance in predicting an individual’s vulnerability to myocardial infarction (MI) through the analysis of a wide range of crucial factors, including demographic and societal standing, medical history, and lifestyle preferences, is noteworthy. This achievement holds the promise of facilitating prompt interventions. Additionally, it could lead to tailored healthcare methodologies intended to reduce potential hazards.

The study is summarized in **Fig 1**. included an analysis of responses gathered by a thorough telephone survey, which covered a variety of subjects including medical history, lifestyle, food habits, and health practices of different individuals. These data were considered for model training and evaluation with an emphasis on myocardial infarction (MI) probability prediction. Given the survey method’s inherent noise, a thorough preprocessing has been performed, including missing data and outliers handling and balancing the training dataset through Minority-weighted Sampling. To optimize the outcomes of training and evaluation, a wide range of machine learning and deep learning models have been applied across different over-sampled ratios which led to the production of a lightweight Artificial Neural Network (ANN) model that ensures good specificity and sensitivity. The model’s prediction reasoning has been deciphered using explainable AI (XAI) approaches, and the individual contributions of each input domain to the predictive framework have been further demonstrated through sensitivity analysis.

**Figure 1.**
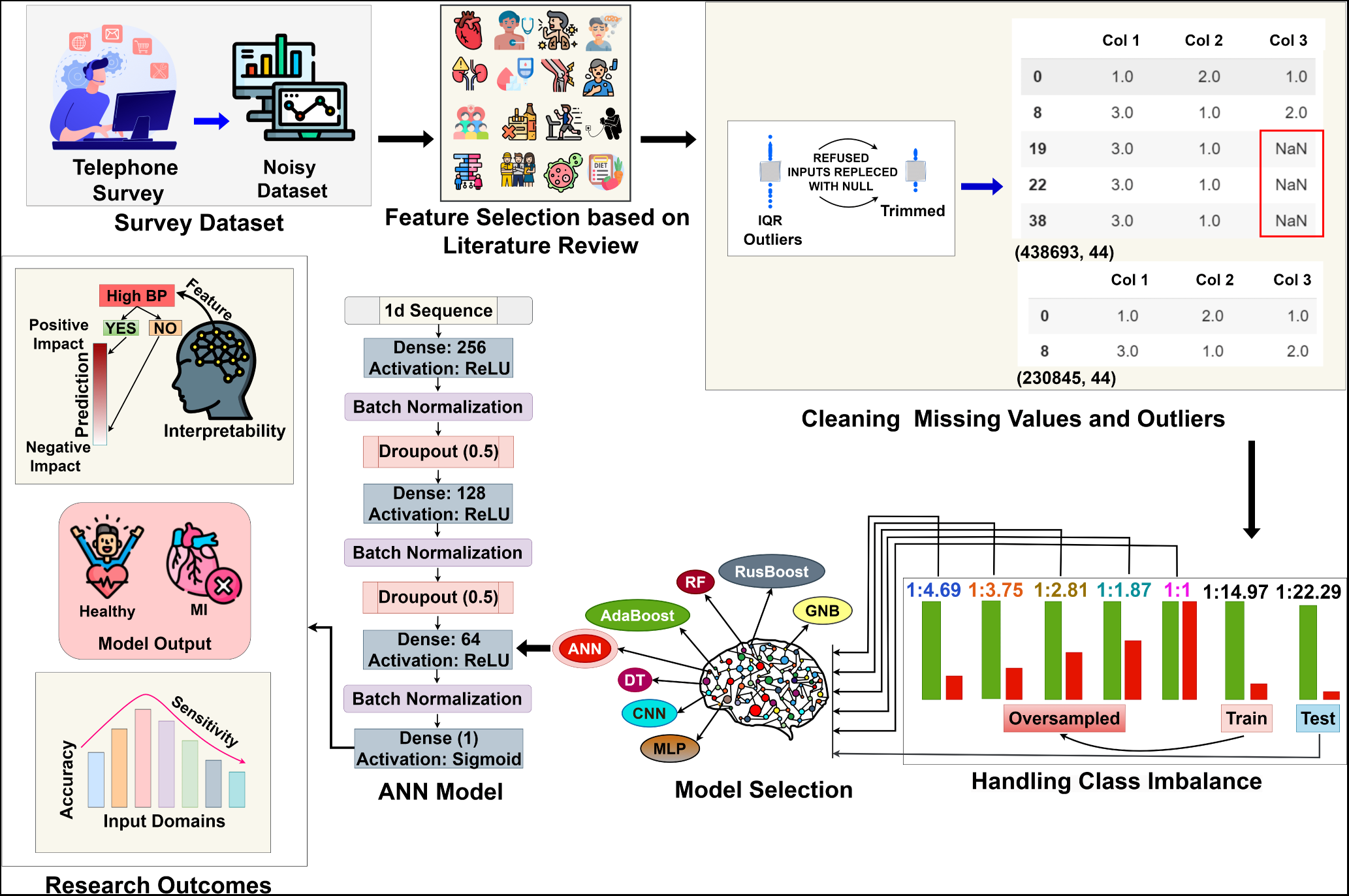
The complete workflow of the myocardial infarction (MI) probability prediction from medical survey data. The methodologies behind developing an ANN model with interpretability to predict myocardial infarction (MI) probability are discussed in this section.

## Feature selection

The study of data from the Behavioural Risk Factor Surveillance System (BRFSS) included a thorough examination of forty-three selected features. These variables have been carefully chosen based on the findings of the research in similar fields^26,27,28,29,31,32,58^. This study provides a broad understanding of the many facets of behavioral risk factors and any potential effects these may have on public health. The dataset was analyzed in order to provide significant insights into the health behaviors and habits of the individuals in terms of myocardial infarction (MI). The variables chosen were dispersed throughout seven various domains in order to obtain a comprehensive picture of health-related behaviors.

Each domain is shown in **Table 2**. concentrated on a specific subject, such as lifestyle choices, the incidence of chronic diseases, indications of mental health, access to treatment, and other crucial elements affecting public health outcomes. This comprehensive study provides useful insights into the interactions between these 43 variables across several domains, contributing to a better understanding of public health trends and factors impacting myocardial infarction (MI).

**Table 2.** Selected features from specific input domains. Forty-three features have been selected from seven different input domains through an in-depth analysis of the dataset to predict myocardial infarction (MI) instances.

| Input Domain | Feature Name |
| --- | --- |
| Socio-demographic factors | Marital status, Residential status, Military record, Reported height, Ethnicity (Race), State, Spoken languages, Gender, Children count, and Age group |
| Economic and social status | Education, Employment status, Earning level (Income), and Mobile usage |
| Medical conditions | Skin cancer history, Other cancer history, Chronic bronchitis history, Heart disease history, Depressive disorder history, Renal disease history (Kidney issues), Diabetes history, Arthritis history, Stroke history, Asthma history, Cholesterol level, Blood pressure level, Self-rated Health status, and BMI level |
| Impairment status | Difficulty seeing, Difficulty dressing or bathing, Difficulty walking, and Difficulty doing errands alone |
| Healthcare availability and affordability | Health insurance, Doctor affordability, Routine check-ups, and Availability of personal health care assistant |
| Personal health habits | Alcohol consumption, Tobacco consumption, Smoking level, Exercise level, and Fruit consumption |
| Preventive health services | HIV status and Vaccination records |

Comprehensive statistical studies are necessary to fully explore the connections between every single input column and the target variable, which is the occurrence of myocardial infarction (MI) cases. It ought to be feasible to determine if there are statistically significant trends or connections between the input variables and the incidence of MI cases by using the proper statistical tests. By assisting in the identification of critical elements or variables that might impact or contribute to the incidence of MI cases, such analyses will provide a deeper comprehension of the fundamental connections between the target variable and the input columns.

In the context of a chi-squared test, the chi-squared statistic (*χ*^2^) is calculated as follows:

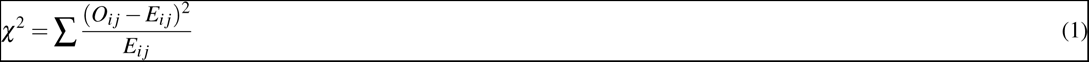

Where: - *O_ij_* represents the observed frequency in a specific cell of the contingency table. - *E_ij_* represents the expected frequency in the same cell. - The summation (∑) is taken over all cells in the contingency table.

The difference between observed and predicted frequencies is quantified by the chi-squared statistic, which gives an indication of the level of correlation between categorical variables.

The p-value (*p*) in the context of a chi-squared test is computed as follows:

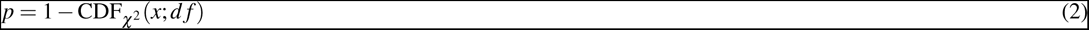

Where: - *x* is the observed chi-squared statistic. - *d f* represents the degrees of freedom, typically calculated as (*r* − 1) × (*c* − 1) for a contingency table with *r* rows and *c* columns.

The p-value is the probability that a chi-squared statistic will be observed that is as extreme to or more extreme than the observed result under the null hypothesis. Stronger statistical significance is shown by a decreased p-value, which implies a relationship between the variables under the analysis.

The level of connection or independence between two categorical variables could be determined by calculating the chi-squared statistic and p-value^59,60^ showed in **Table 3**. The chi-squared statistic is a measure used to evaluate the difference between actual and predicted data frequencies in a contingency table. A greater correlation between the variables is indicated by a bigger chi-squared value. The p-value shows the chance of a random connection, a lower p-value (below 0.05) means a stronger, more statistically significant relationship between variables. The relevance of input features differs when it comes to the examination of the input domains. A range for p-values and chi-squared values is conducted for the socio-demographic domain, suggesting both strong and weak associations with the target column. The area of economic and social status, on the other hand, exhibits robust correlations with lower p-values and a wider range of chi-squared values. The impairment status domain likewise shows substantial links to the target, while the medical conditions domain includes several input variables with extremely high connections to the target column. On the other hand, although the low p-values imply some degree of relevance, preventive health services, and personal health practices have the least chi-squared range, indicating lower correlations with the target column.

**Table 3.** Statistical measures for the specified input domains to establish the relationship between these domains and the target variables. An increased discrepancy between the observed and predicted frequencies is shown by a bigger chi-squared value, which implies a stronger correlation between the input variables and the target. A lower p-value suggests that there is a greater likelihood of the observed correlation being the result of chance, which increases its statistical significance.

| Input Domain | Chi-squared Range | P-value Range |
| --- | --- | --- |
| Socio-demographic factors | 5.24 - 5945.99 | 0.0 - $2.72 \times 10^{-94}$ |
| Economic and social status | 699.12 - 6327.68 | 0.0 - $9.46 \times 10^{-148}$ |
| Medical conditions | 130.38 - 47095.41 | 0.0 - $9.17 \times 10^{-56}$ |
| Impairment status | 1326.34 - 5126.42 | 0.0 - $2.13 \times 10^{-290}$ |
| Healthcare availability and affordability | 57.86 - 4738.84 | 0.0 - $4.25 \times 10^{-198}$ |
| Personal health habits | 18.35 - 2263.56 | 0.0 - $2.26 \times 10^{-43}$ |
| Preventive health services | 66.26 - 250.47 | $2.05 \times 10^{-56}$ - $3.96 \times 10^{-16}$ |

## Data preprocessing

The data collection process carried out via telephone surveys, has produced a significant number of missing values and outliers. Additionally, the dataset has a class imbalance issue. Despite these issues, this large dataset remains a great resource for learning about myocardial infarction (MI) prevalence and risk factors. To guarantee the robustness and reliability of the study, appropriate methods for preprocessing the data are required. A variety of preprocessing steps were methodically used on the dataset with selected features detailed in **Fig 1**. The Interquartile Range (IQR) was used to identify the existence of outliers in these features. In the survey, many individuals refused to provide answers or didn’t know the exact answers at that time. In the dataset, 292991 rows have been identified as containing refused inputs. These incidents were identified by putting specific input values in the dataset. This study highlights most of the identified outliers in the dataset are the causes of these refusal responses from individuals. To trim the outliers from the dataset, a custom function has been introduced that can identify invalid responses from the specific features, and replace these values as null values. Accordingly, all identified null values were removed from the dataset. The target column was encoded to make it compatible with the AI algorithms. To aid in the training and evaluation of artificial intelligence (AI) models, the dataset was split with a split ratio of 51.14% for training and 48.86% for testing. This distribution was selected to obtain the best results from uneven data. A substantial class imbalance issue was found in the dataset, where the data for healthy individuals vastly outnumbered those for myocardial infarction (MI) cases. There were initially 110659 instances of healthy cases and 7393 instances of myocardial infarction (MI) cases in the trainset. Similarly, the testset included 107829 healthy cases and 4964 cases of myocardial infarction (MI). In order to effectively extract findings from the imbalanced data, data for the minority class was carefully balanced through Minority-weighted Sampling in order to address this variation in model training. The outcomes from different over-sampled ratios across several models were compared to identify the ideal ratio for model training. The categories and values of each specified domain such as socio-demographic, economic and social, medical condition, impairment status, healthcare availability and affordability, personal health habit, and preventive health service are presented respectively in the appendix in **Table 6**, **Table 7**, **Table 8**, **Table 9**, **Table 10**, **Table 11**, and **Table 12**.

## Sensitivity analysis

The preprocessed dataset covers information from seven distinct input domains, including socio-demographic factors, economic and social status, medical conditions, impairment status, healthcare availability and affordability, personal health habits, and preventive health services. Data from these domains were collected from the individuals, and it is critical to validate the model’s sensitivity to each of these domains thoroughly. To do this, each input domain was used individually as training data for the prediction model, and the resulting performances were carefully observed for each scenario. The proposed model is then trained on the training sets for the selected domains, and the accuracy for each domain is computed. However, considering concerns with class imbalance, accuracy alone is not regarded as a meaningful performance metric. Following, recall of both target classes is employed for each data domain. Similarly, the Area Under the Curves (AUC) and scores are also generated for all selected domains in the next part. Finally, SHapley Additive exPlanations (SHAP) values are used to measure the relevance of features from each domain, with higher SHAP values indicating a greater impact on the model’s performance. This work intends to obtain insights into the model’s robustness, generalization capabilities, and potential biases by systematically evaluating its performance across multiple input domains, improving overall understanding and reliability in its application across a variety of scenarios.

### 0.1 Analyzing accuracy from specified domains

Data from individuals across different areas were used to measure its accuracy in various domains. The study evaluated how well the model performed in predicting outcomes based on different domains. If a specific data domain is chosen for myocardial infarction (MI) probability prediction, the accuracy of the prediction for the selected data within that domain is analyzed in this section.

In this thorough study depicted in **Fig 2**, data of individuals from various subject areas accumulated, and the evaluated model’s accuracy across each domain was calculated through the proposed Artificial Neural Networks (ANN) model. The description of the variables in each domain is included in **Table 2**. The domain that was relevant to medical records performed noticeably and possessed the highest accuracy, which was the expected outcome of this study. However, impairment status which contains information about individuals having disabilities in doing regular work surprisingly performed remarkably well and possessed the second-highest accuracy. Accordingly, the domains related to economic and social status, healthcare availability and affordability, and socio-demographic characteristics are identified to have moderate contributions towards the prediction. Personal health habits and preventive health services domains showed the least accuracy, which indicates a minimal contribution to the prediction. This method has enabled the facts to distinguish the various contributions of different domains to the model’s predictions, revealing insight into which domains have the most influence on the model’s performance. Due to the imbalanced class distribution, assessing recall for both target classes is essential for a more meaningful evaluation of performance, as only accuracy can be misleading in this case. Determining the true positive rate as well as the true negative rate is essential to extract more important and informative insights about the input domains which is a primary emphasis in this study. These metrics provide a more detailed view of the model’s performance by illuminating the model’s capacity to accurately identify instances that are positive (true positives) and instances that are negative (true negatives).

**Figure 2.**
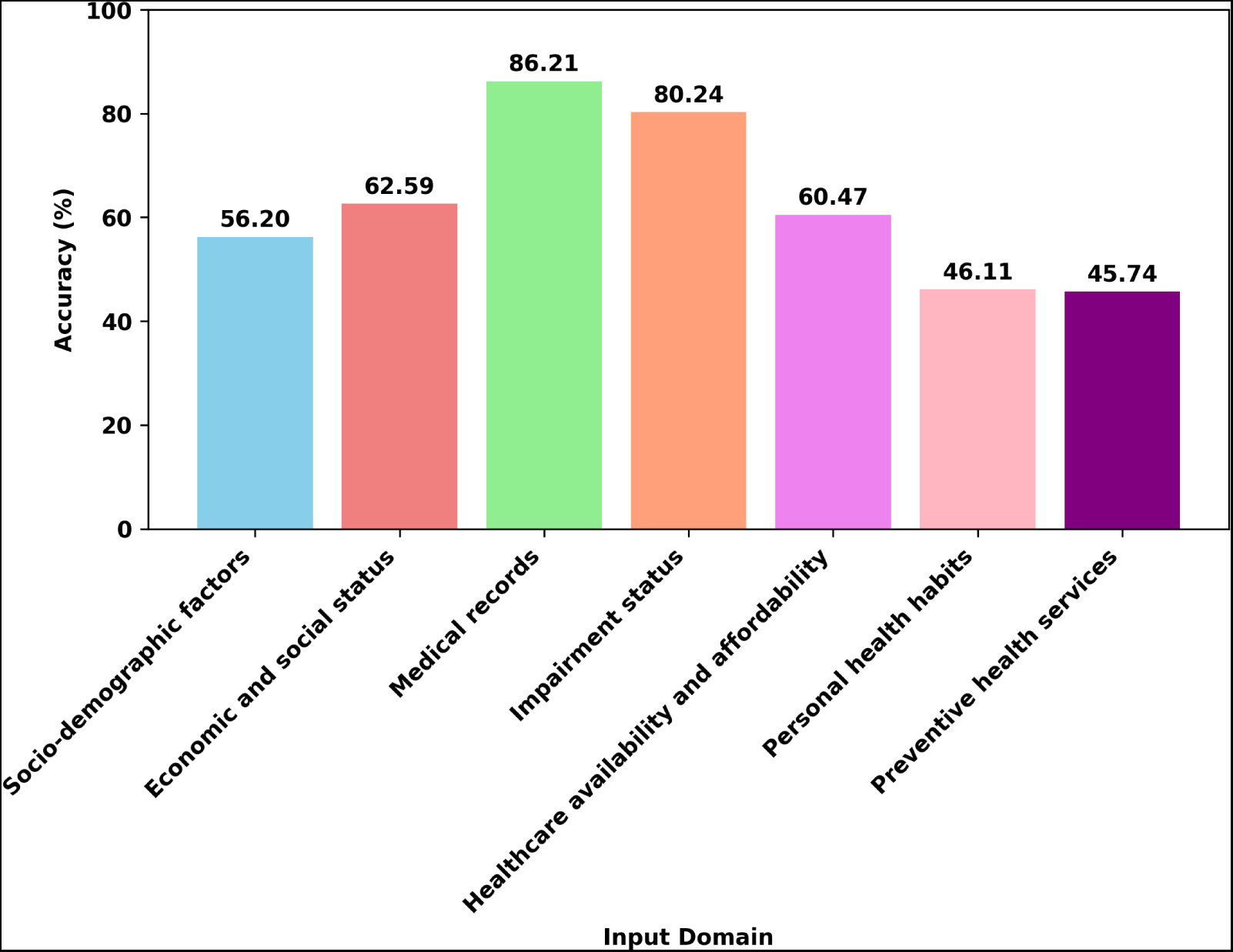
Sensitivity of the proposed model towards each input domain. Separate input domains were used as individual train and test sets for model learning and evaluation. The resulting accuracies for each scenario are shown in this section. Each bar is colored differently to show accuracies for a distinct input domain.

### 0.2 Analyzing recall from specified domains

Data from individuals across different areas were used to measure the true positive rates for classifying healthy cases and myocardial infarction (MI) cases in distinct domains. The study evaluated how well the model performed in predicting both target classes based on different domains. If a specific data domain is chosen for myocardial infarction (MI) probability prediction, the recall of both target classes for the selected data is analyzed in this section.

In this thorough study depicted in **Table 4**, the evaluated model’s true positive rates of the class 0 (healthy cases) and class 1 (MI cases) across each domain was conducted through the proposed Artificial Neural Networks (ANN) model. The recall rates for the healthy cases range from 0.55 to 0.88 across the specified domains, illustrating the model’s efficiency in properly recognizing healthy instances within each domain. The MI cases recall rates, on the other hand, range from 0.42 to 0.79, suggesting the model’s ability to diagnose myocardial infarction reliably. The socio-demographic factors along with economic and social status domains obtained relatively higher recall scores for MI cases compared to healthy cases. However, the socio-demographic factors domain indicates a high recall imbalance for healthy and MI cases. The medical conditions domain obtained a high recall for healthy cases and a comparatively standard recall for MI cases. The impairment status as well as healthcare availability and affordability domains obtained unbalanced recall ratios for healthy and MI cases. The personal health habits and preventive health services domains obtained comparatively lower recall scores for both cases. This thorough breakdown not only illustrates the model’s individual strengths and weaknesses within each input area but also emphasizes the varied degrees to which these domains impact prediction performance. These insights are crucial for evaluating the model’s performance and adapting its applicability to various scenarios and circumstances.

**Table 4.**
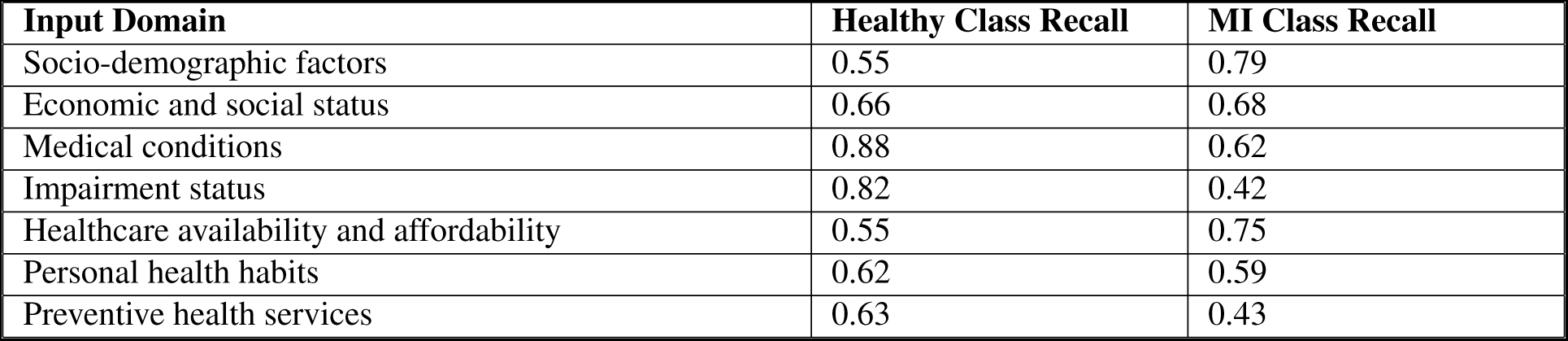
True positive rates of the class 0 (healthy cases) and class 1 (MI cases) for the selected features from specific input domains using the proposed Artificial Neural Networks (ANN) model to state the significance of the input domains in the prediction. Individual train and test sets were created from separate input domains for model development and evaluation. This section depicts the recall obtained for each circumstance.

### 0.3 Analyzing Area Under the Curve (AUC) from specified domains

Data from individuals across different areas were used to measure the Area Under the Curve (AUC) for predicting myocardial infarction (MI) probability in distinct domains. The study evaluated how well the model performed in predicting true positives and false positives rates based on different domains. If a specific data domain is chosen for myocardial infarction (MI) probability prediction, the AUC of the prediction for the selected data is analyzed in this section.

The accompanying **Fig 3** depicts the obtained true positive and false positive rates for the selected features from specific input domains. The AUC assessments for the different classification models in the scenarios offer insight into performance in terms of true positive rate and true negative rate. The socio-demographic factors, economic and social status, as well as healthcare availability and affordability domain, obtained relatively higher AUC scores. The medical conditions domain obtained a maximum AUC score. The personal health habits, impairment status, and preventive health services domain obtained a comparatively lower AUC score. The observed variances in the performance of different domains while assessing the predictive performance, as evaluated by the Area Under the Receiver Operating Characteristic Curve (AUC), were examined in this detailed analysis. The demographics, economic and social status, and healthcare accessibility and affordability domains, in particular, exhibited considerably higher AUC scores, demonstrating usefulness in predicting important outcomes. Notably, the domain of medical conditions possessed the highest AUC score, demonstrating its excellent predictive potential. Personal health behaviors, impairment status, and preventive health services domains, on the other hand, had lower AUC values, indicating a reduced potential in relative prediction. These findings shed light on the relative strengths and weaknesses of the various areas examined in this study.

**Figure 3.**
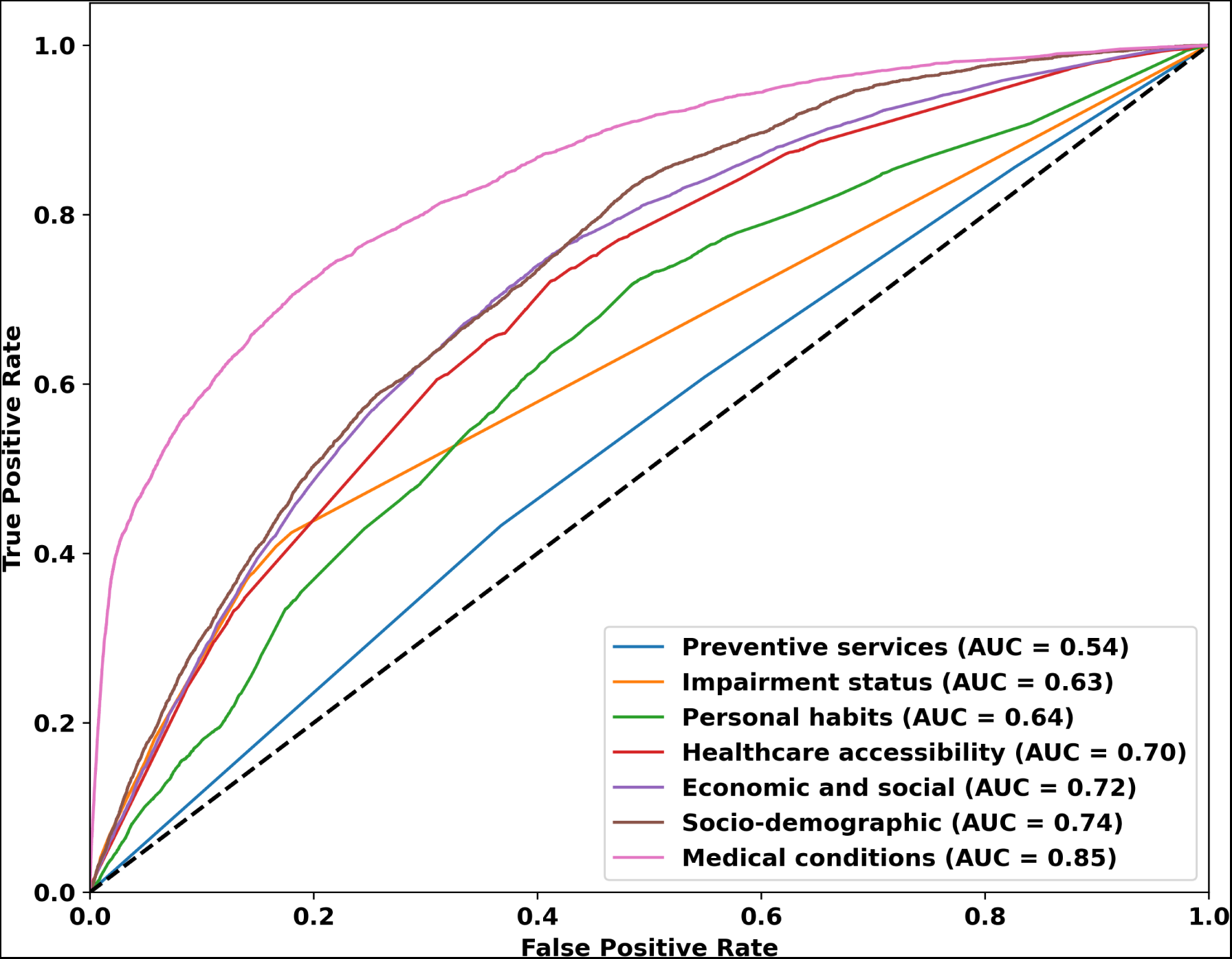
Area Under the Receiver Operating Characteristic (ROC) curve for the selected features from specific input domains using the proposed Artificial Neural Networks (ANN) model. AUC curves for each input domain are implemented in this section to demonstrate the true positive and false positive rates.

### 0.4 Analyzing most impactful features from specified domains

An analysis of SHapley Additive exPlanations (SHAP) values was employed to estimate the importance of features in prediction. SHAP values evaluate feature importance by evaluating each feature’s contribution to the difference between the model’s anticipated result and the average prediction. This ensures that credit is distributed fairly among features. The analysis is conducted by assigning different shades for different domains demonstrating the importance of each feature within specified input domains towards predictions. This provides an overview of which features from particular domains contribute more to predictions than features from other domains. The approach provides insight into the relative relevance of features and the domains in determining model predictions.

The study is critical in understanding the risk factors for myocardial infarction (MI), adopting a comprehensive strategy that determines the relevance of each feature, characterized by a spectrum of colors to represent distinct input domains detailed in **Fig 4**. Notably, the data show that variables from the medical condition domain showed the greatest influence on predicting myocardial infarction (MI), highlighting the importance of diverse health-related issues and individual medical conditions on prediction performance. Furthermore, socio-demographic aspects appear as major drivers, illuminating the complex relationship between an individual’s social and demographic circumstances and predictive performance. Equally, characteristics connected with economic and social standing have a significant impact, emphasizing the critical role of educational and financial situations in determining the prediction. Features anchored in the impairment status domain stand out as well, emphasizing the factors of an individual’s mobility and visual impairments affecting the predicted outcome. Furthermore, the existence and accessibility of healthcare resources, as defined within the area of healthcare availability and cost, play a significant impact, emphasizing the influence of medical aid and doctor availability on prediction performance. Furthermore, the study emphasizes the significant effect of personal health behaviors and areas of preventive health services, highlighting the deep impact of an individual’s lifestyle choices and sincerity regarding health on the prediction model. Only recognizing the influence of the input features is insufficient to establish the meaningfulness of these inter-domain feature impacts. As a result, the study emphasizes the vital requirement for interpretability in order to extract and identify useful insights, hence increasing the study’s overall significance and relevance, which is one of the study’s key objectives.

**Figure 4.**
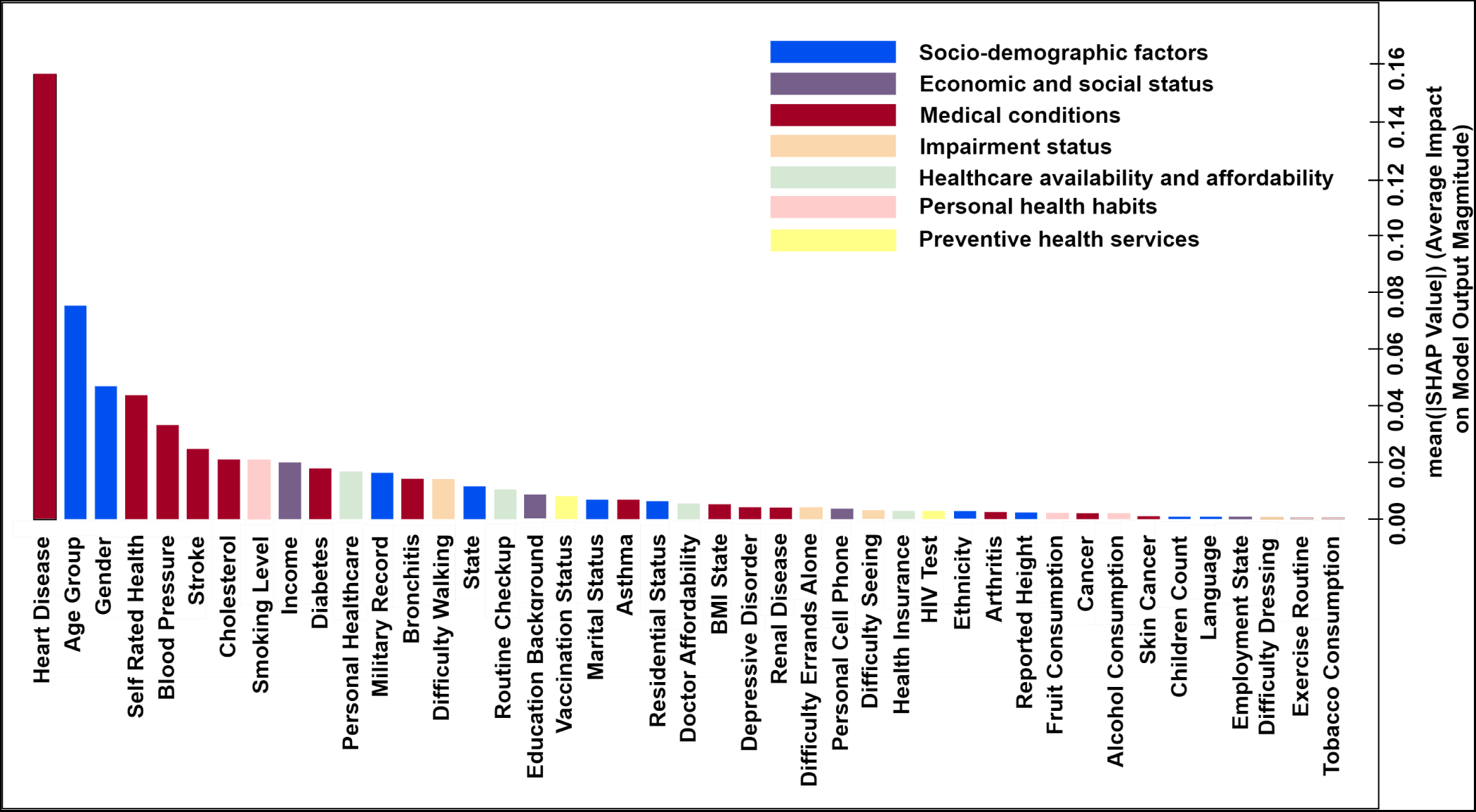
Feature importance employing SHapley Additive exPlanations (SHAP) values using the proposed Artificial Neural Networks (ANN) model. The significance of each feature from specified input domains toward prediction was shown in this section. Each color shade represents an input domain. The features are listed in descending order of importance.

## Interpretability analysis

In the context of medical platforms, explainable AI (XAI) is crucial^61^, particularly when it comes to the prediction of myocardial infarction (MI) probability using survey data. Transparency and interpretability^62^ are crucial in the healthcare industry since decisions based on AI-driven models may have far-reaching effects^32,34^. In addition to improving the credibility and dependability of predictive models, XAI provides healthcare professionals with the knowledge required to comprehend the logic behind AI-generated predictions. The development of more precise and reliable strategies for the early detection of myocardial infarction (MI), improving patient outcomes and ensuring the responsible deployment of AI in healthcare are all enhanced by this understanding, which also promotes informed decision-making. The analysis has been conducted in this study using data from multiple domains. The socio-demographic domain contains categories such as marital status, residential status, military record, reported height, ethnicity (race), state, spoken languages, gender, children count, and age group. Following, the economic and social status domain contains categories such as education, employment status, earning level (income), and mobile usage. Accordingly, the medical conditions domain is represented by a diverse set of categories, encompassing skin cancer history, other cancer histories, chronic bronchitis history, heart disease history, depressive disorder history, renal disease history (kidney issues), diabetes history, arthritis history, stroke history, asthma history, cholesterol level, blood pressure level, self-rated health status, and BMI level. Consecutively, the impairment status domain comprises categories related to difficulty in seeing, dressing or bathing, walking, and doing errands alone. Accordingly, the healthcare availability and affordability domain includes categories of health insurance, doctor affordability, routine checkups, and the availability of a personal healthcare assistant. Following, the personal health habits domain contains categories such as alcohol consumption, tobacco consumption, smoking level, exercise level, and fruit consumption. Finally, the preventive health services domain comprises categories by HIV status and vaccination records.

One method of understanding ML predictions is to computer and interpret SHapley Additive exPlanations (SHAP) values. SHAP aims to determine precisely how much each of the features of the model contributes to the ultimate outcome. It does so by generating a weighted average of the contribution of each of its features, attempting to account for every possible combination^42^. The SHAP values of this proposed model have been analyzed in this study to assess the impact of each feature on a model’s prediction by assigning a contribution value to each feature, helping to understand the individual effects. Following, the features that have significant contributions in predicting myocardial infarction (MI) are grouped by domains then the contribution of each category inside a feature has been analyzed to assess the individual contribution. This provides a clear picture of how each category inside features from a specific domain influence the myocardial infarction (MI) and highlights dependencies between the myocardial infarction (MI) and domain-specific input features.

### 0.5 Analyzing feature effects

Features from seven data domains were considered as input in this study to predict the probability of myocardial infarction (MI). To identify which features from a specified domain have the most impact on predicting myocardial infarction (MI) cases, the effect of each feature on this has been analyzed using the SHapley Additive exPlanations (SHAP) values. SHAP works as the hiring detectives for each of these features. Each detective checks how much the prediction changes when one feature is considered or not. It’s like shuffling the input features to observe different possibilities. After exploring all options, SHAP reveals which input features have the most impact on predicting the chance of a myocardial infarction (MI) and how much each matters.

The SHAP summary graphic detailed in **Fig 5**, which is ranked by importance, and demonstrates the way different features affect the model’s prediction. The X-axis represents SHAP values for a specific feature. Positive SHAP values show a feature’s contribution to the prediction for the class 1 (MI cases). Negative SHAP values imply that the presence or higher values of that feature contribute to the model predicting class 0 (healthy cases) rather than class 1 (MI cases). When a feature’s SHAP value is nearly 0, it indicates that the feature has little to no influence on the model’s prediction in that specific instance. The Y-axis represents feature distributions. The prediction is most significantly increased by the top features, whereas the bottom features have a smaller impact. The specified features have multiple categories represented by numbers. In the color bar, red denotes greater input values, and blue denotes lower input values. Each input category inside features represents a special group of individuals. Hence, the graphic shows which special groups have a significant impact on a specific target class. The categories in each input feature along with data frequencies are presented in the appendix in **Table 6**, **Table 7**, **Table 8**, **Table 9**, **Table 10**, **Table 11**, and **Table 12**.

**Figure 5.**
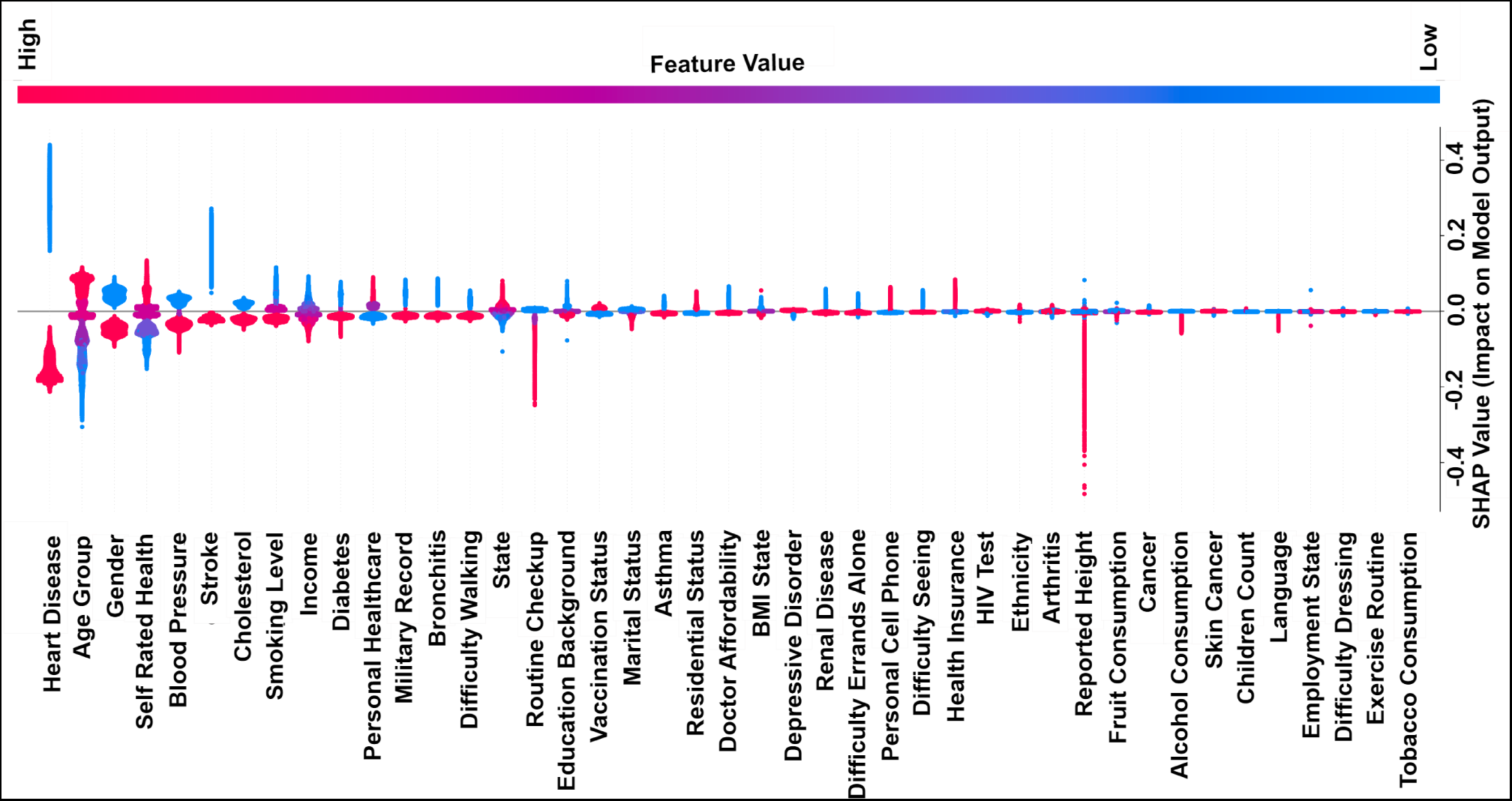
A summary graphic that employs SHapley Additive exPlanations (SHAP) values to combine feature importance and feature effects of the selected features from the preprocessed dataset. The graphic is positioned in a rotated manner. Each point on the x-axis indicates a Shapley value for a particular feature. When it comes to Shapley values, higher values indicate the most significant features and lower values denote fewer significant features. The features are listed in descending order of importance on the y-axis. The color of the dots corresponds to the respective feature categories from the dataset, which range from low to high. Each feature category represents a specific group of individuals.

The observations obtained using SHAP regarding input categories that have a significant impact on target class 1 (MI cases) mentioned in **Fig 5**, are discussed in this section. If certain categories from the input claimed to have a positive impact on class 1 (MI cases) if and only if that specific category has positive SHAP values. Individuals with characteristics from the medical condition domain such as heart disease history, high blood pressure, stroke history, high cholesterol, diabetes history, bronchitis, and renal disease history possess a significant contribution to the prediction. Similarly, individuals with reported bad health conditions possess a significant contribution to the prediction. Further, individuals with skin cancer or any other cancer history possess a minimal contribution to the prediction. In a similar vein, individuals who are underweight and overweight possess comparatively more contributions to the prediction. Accordingly, individuals from the impairment domain with characteristics such as difficulty walking or climbing, difficulty doing errands alone, and disability seeing possess a significant contribution to the prediction. Likewise, individuals having difficulty dressing or bathing possess minimal contributions to the prediction. In addition, individuals from the economic and social status domain with characteristics such as low income and less education possess a significant contribution to the prediction. Besides, it can be said individuals who use significantly more phones and individuals who don’t have cell phones, these two groups possess comparative contributions to the prediction. Further, individuals who work for wages possessed minimal contributions to the prediction. Next, individuals from the healthcare availability and affordability domain with characteristics such as individuals who have no personal healthcare provider or who have more than one personal healthcare provider, individuals who faced financial barriers to afford a doctor in need in the past year, and individuals who have no coverage of health insurance possessed considerable contributions to the prediction. Besides, the routine checkup schedule duration of individuals has a minimal contribution to the prediction. Furthermore, individuals from the social-demographic factors domain with characteristics such as the elderly and senior adults group, males, individuals who have a military service record, individuals living in the states associated such as Guam, Puerto Rico, and the Virgin Islands, individuals who are married, individuals who don’t have proper living arrangements, and individuals who belong to minor non-Hispanics possessed comparative contributions to the prediction. Accordingly, the reported height of the individuals and individuals with more children possessed minimal contributions to the prediction. Likewise, individuals from the personal health habits domain with characteristics such as individuals who have smoking habits particularly chain smokers or regular tobacco consumers possessed a significant contribution to the prediction. However, characteristics from the preventive health services domain seem to have no significant observation related to the prediction.

### 0.6 Analyzing effects and dependencies

The input features that were used in this study have multiple categories. The effect of these features on predicting myocardial infarction (MI) cases has been analyzed earlier in **Fig 5**. However, to analyze how much each category inside these features from the different domains contributes to predicting myocardial infarction (MI) cases have been analyzed in this section using the SHapley Additive exPlanations (SHAP) values. The features that have been identified as important for predicting myocardial infarction (MI) cases in **Fig 5**, the categories inside these selective features have been analyzed in this section to measure the impact of each category inside these features in predicting myocardial infarction (MI) probability using SHAP dependencies. A SHAP dependency shows how the output of a model depends on the values of a specific feature, helping to understand the impact of that feature on the model’s predictions and how relevantly that feature interacts with another feature. The motive of this analysis is to identify the key risk factors for myocardial infarction (MI).

#### 0.6.1 Analyzing effects and dependencies from medical condition domain

The effects and dependencies of each category inside the features from the medical condition domain that have an impact on predicting myocardial infarction (MI) cases have been analyzed in this section. This analysis also shows how these selective features from the medical condition domain interact with the other features used in this study to influence the prediction of myocardial infarction (MI) probability.

This section shown in **Fig 6** provides a detailed explanation regarding the way features of the medical condition domain are contributed to predicting MI cases, including the manner in which interact with other features. The x-axis represents the categories inside specific features in the medical condition domain. The categories in each medical condition input feature along with data frequencies are discussed in the appendix in **Table 6**. The y-axis shows the associated SHapley Additive exPlanations values for each category in the same feature of the medical condition domain. The color shades in the color bar represent input categories of different features that interact with the examined feature. Each color shade in the graphic represents a complicated interaction effect between the medical condition and other specified attributes including general health, age, and heart disease. This provides a clear understanding of the intricate interplay of factors associated with the medical condition domain that affects the model’s output.

**Figure 6.**
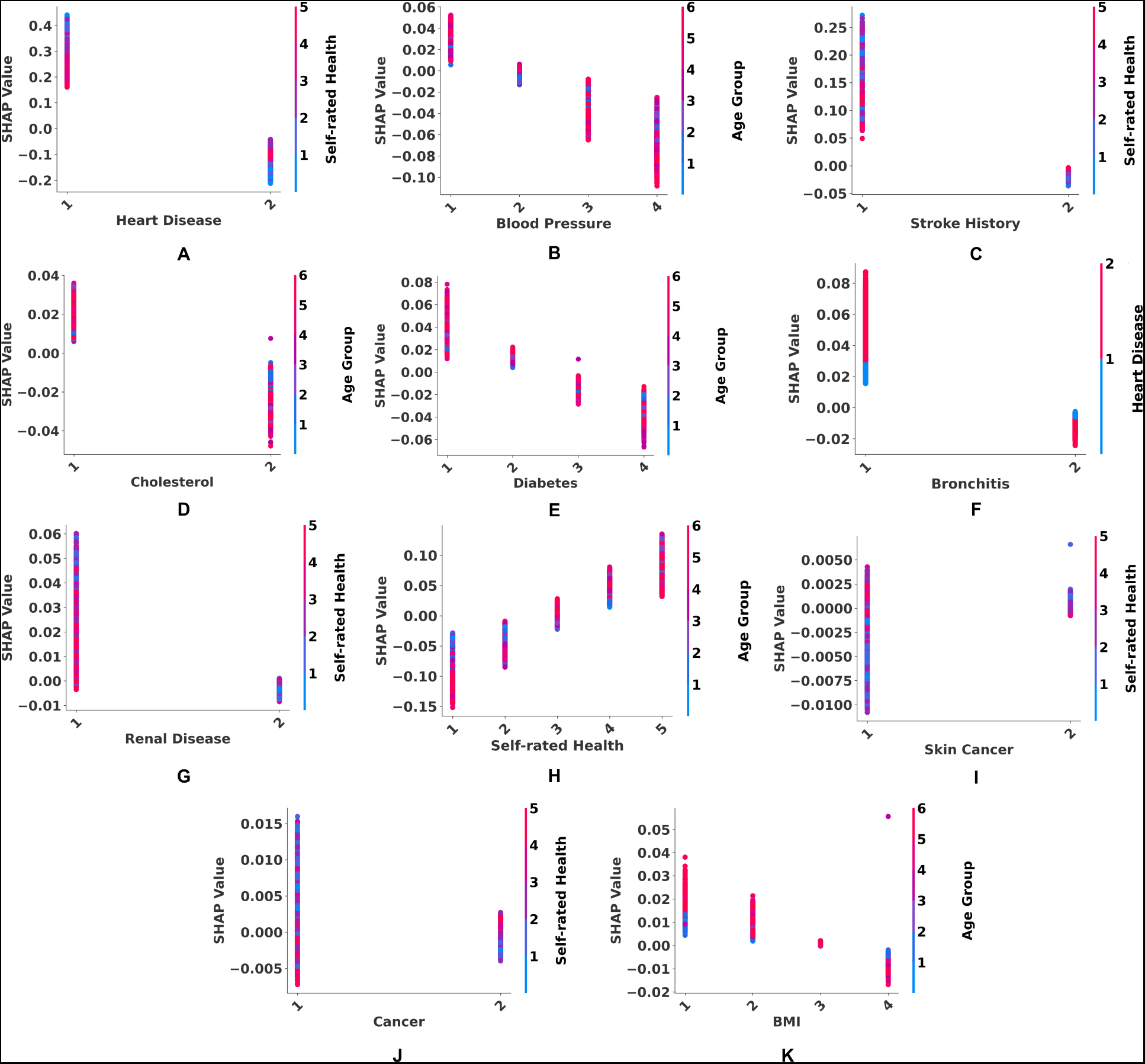
A dependence graphic to represent the predictive effect of each category in medical condition features with interaction effects. SHapley Additive exPlanations (SHAP) values were employed to represent the effect of each category in medical condition in terms of identifying myocardial infarction (MI) probability including the interaction effects with other features. The x-axis represents the different categories inside a specific feature of a medical condition. Besides, the y-axis represents SHAP values for an input category inside that specific feature. The color bar represents different input categories from another feature that interacts with that specific feature.

Identifying health outcomes associated with medical conditions by predictive modeling requires careful evaluation of several interacting factors. Each interaction between medical conditions and other specified attributes including general health, age, and heart disease claimed to have an impact on predicting MI cases if and only if it has positive SHAP values. The **Fig 6** in part **A** shows how general health conditions affect the heart disease individuals to have a probability of having MI. Individuals who have heart diseases, especially those with reported poor health statuses, have a stronger impact on the prediction. In a similar vein, the **Fig 6** in part **B** shows elderly individuals with hypertension are more significant in the prediction model than youths with hypertension. Interestingly, the **Fig 6** in part **C** shows how general health conditions affect the individuals with a stroke history to have a probability of having MI. Individuals with a history of strokes, especially those with reported poor health statuses, have a stronger impact on the prediction. Moreover, the **Fig 6** in part **D** shows high cholesterol has a stronger predictive effect on elderly individuals than on young. Additionally, the **Fig 6** in part **E** shows the impact of diabetes on predictions tends to lean towards the elderly, since the disease has a greater predictive effect than it does on younger individuals. Surprisingly, the **Fig 6** in part **F** shows individuals with bronchitis have a stronger influence on predictions. Besides, individuals with both bronchitis and heart disease have a significant influence on the predictions. Furthermore, **Fig 6** in part **G** shows those with renal issues and reported to be in poor health have a stronger impact on the prediction. Accordingly, the **Fig 6** in part **H** shows elderly individuals with reported poor health have a greater influence on prediction than younger individuals with similar reports of poor health. At the very least, the **Fig 6** in part **I** and **J** shows those with a history of skin or other cancers, especially those who claim poor health have a stronger impact on the prediction. Remarkably, the **Fig 6** in part **K** shows elderly individuals with low BMI contribute more to the predictions than youths with low BMI, and even those who are underweight make a stronger contribution to the prediction than those who are overweight.

#### 0.6.2 Analyzing effects and dependencies from impairment status domain

The effects and dependencies of each category inside the features from the impairment status domain that have an impact on predicting myocardial infarction (MI) cases have been analyzed in this section. This analysis also shows how these selective features from the impairment status domain interact with the other features used in this study to influence the prediction of myocardial infarction (MI) probability.

This section shown in **Fig 7** provides a detailed explanation regarding the way features of the impairment status domain are contributed in predicting MI cases, including the manner in which interact with other features. The x-axis represents the categories inside specific features of the impairment status domain. The categories in each impairment status input feature along with data frequencies are discussed in the appendix in **Table 7**. The y-axis shows the associated SHapley Additive exPlanations values for each category in the same feature of the impairment status domain. The color shades in the color bar represent input categories of different features that interact with the examined feature. Each color shade in the graphic represents a complicated interaction effect between the impairment status and other specified attributes including heart disease and general health. This provides a clear understanding of the intricate interplay of factors associated with the impairment status domain that affects the model’s output.

**Figure 7.**
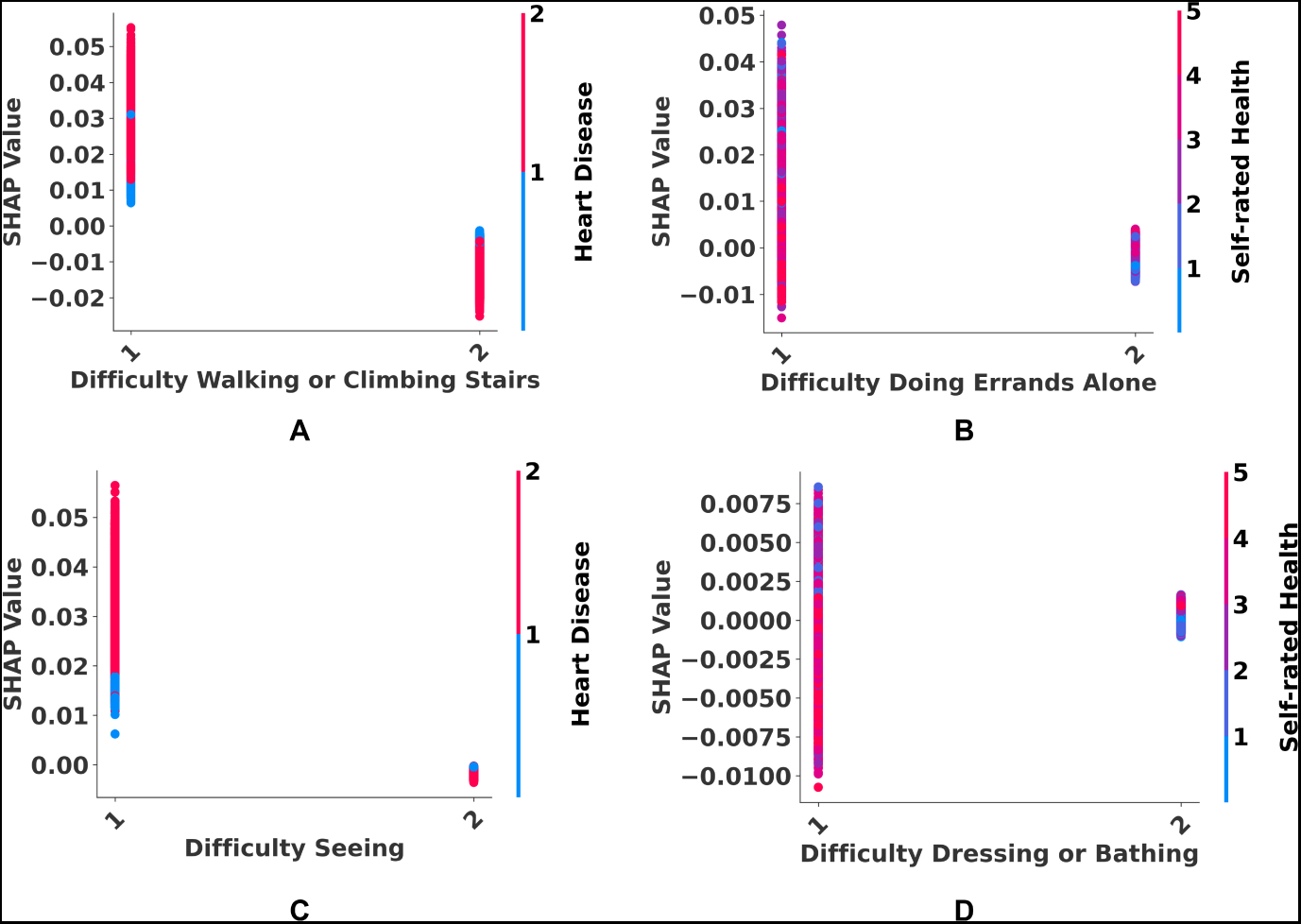
A dependence graphic to represent the predictive effect of each category in impairment status features with interaction effects. SHapley Additive exPlanations (SHAP) values were employed to represent the effect of each category in impairment status in terms of identifying myocardial infarction (MI) probability including the interaction effects with other features. The x-axis represents the different categories inside a specific feature from impairment status. Besides, the y-axis represents SHAP values for an input category inside that specific feature. The color bar represents different input categories from another feature that interacts with that specific feature.

Identifying health outcomes associated with impairment status by predictive modeling requires careful evaluation of several interacting factors. Each interaction between impairment status and other specified attributes including heart disease and general health claimed to have an impact on predicting MI cases if and only if it has positive SHAP values. The **Fig 7** in part **A** shows how heart disease affects individuals with walking or climbing obstacles to have a probability of having MI. Individuals who have walking or climbing obstacles, especially those with heart diseases, have a minimal contribution to the prediction. Furthermore, the **Fig 7** in part **B** shows those who report having trouble running errands and self-reporting being in bad health have a stronger impact on the prediction. In a similar vein, the **Fig 7** in part **C** shows individuals who have vision issues, especially those with heart diseases have a significant impact on the prediction. Lastly, the **Fig 7** in part **D** shows those who struggle with dressing and bathing have a stronger influence on predictions when reporting being in bad health.

#### 0.6.3 Analyzing effects and dependencies from economic and social status domain

The effects and dependencies of each category inside the features from the economic and social status domain that have an impact on predicting myocardial infarction (MI) cases have been analyzed in this section. This analysis also shows how these selective features from the economic and social status domain interact with the other features used in this study to influence the prediction of myocardial infarction (MI) probability.

This section shown in **Fig 8** provides a detailed explanation regarding the way features of the economic and social status domain are contributed to predicting MI cases, including the manner in which interact with other features. The x-axis represents the categories inside specific features of the economic and social status domain. The categories in each economic and social status input feature along with data frequencies are discussed in the appendix in the appendix in **Table 8**. The y-axis shows the associated SHapley Additive exPlanations values for each category in the same feature of the economic and social status domain. The color shades in the color bar represent input categories of different features that interact with the examined feature. Each color shade in the graphic represents a complicated interaction effect between the economic and social status and other specified attributes including age, walking or climbing difficulty, and gender. This provides a clear understanding of the intricate interplay of factors associated with the economic and social status domain that affects the model’s output.

**Figure 8.**
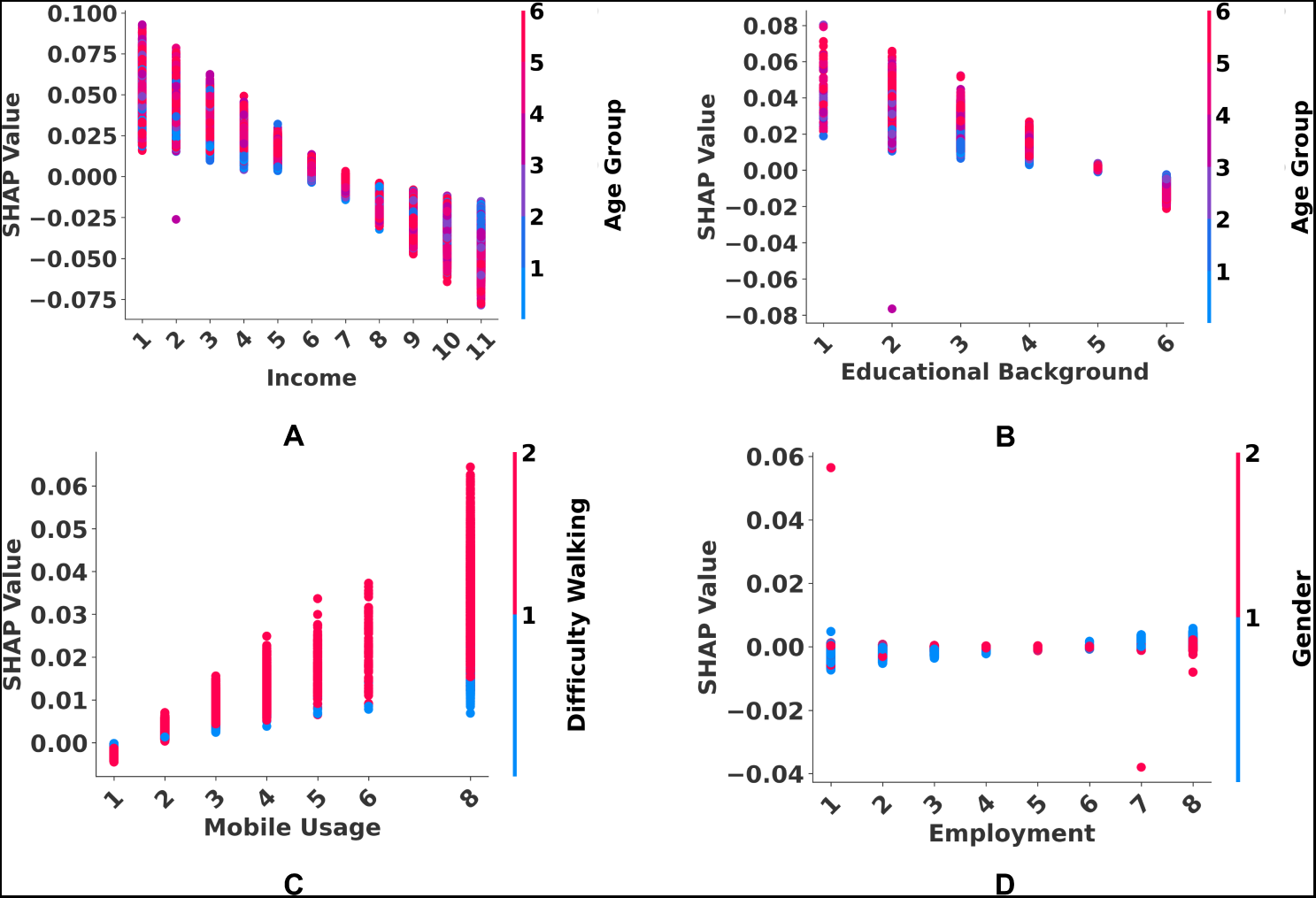
A dependence graphic to represent the predictive effect of each category in economic and social status features with interaction effects. SHapley Additive exPlanations (SHAP) values were employed to represent the effect of each category in economic and social status in terms of identifying myocardial infarction (MI) probability including the interaction effects with other features. The x-axis represents the different categories inside a specific feature from economic and social status. Besides, the y-axis represents SHAP values for an input category inside that specific feature. The color bar represents different input categories from another feature that interacts with that specific feature.

Identifying health outcomes associated with economic and social status by predictive modeling requires careful evaluation of several interacting factors. Each interaction between economic and social status and other specified attributes including age, walking or climbing difficulty, and gender claimed to have an impact on predicting MI cases if and only if it has positive SHAP values. The **Fig 8** in part **A** shows when it comes to elderly individuals, the predictive impact of lower income is significantly stronger than it is for younger individuals with lower incomes. Furthermore, the **Fig 8** in part **B** shows elderly individuals with no or limited educational background have a greater influence on the prediction than younger adults with comparable educational backgrounds. Furthermore, the **Fig 8** in part **C** shows individuals using no cell phone or more cell phones have a greater influence on predictions than individuals. This also interacts with another attribute to show individuals using no cell phone or more cell phones with walking or climbing difficulties have a minimal prediction impact. Moreover, the **Fig 8** in part **D** shows when it comes to employment status, men who are disabled, retired, or working for a wage have significantly higher predictive significance than women in similar situations.

#### 0.6.4 Analyzing effects and dependencies from healthcare availability and affordability domain

The effects and dependencies of each category inside the features from the healthcare availability and affordability domain that have an impact on predicting myocardial infarction (MI) cases have been analyzed in this section. This analysis also shows how these selective features from the healthcare availability and affordability domain interact with the other features used in this study to influence the prediction of myocardial infarction (MI) probability.

This section shown in **Fig 9** provides a detailed explanation regarding the way features of the healthcare availability and affordability domain are contributed to predicting MI cases, including the manner in which interact with other features. The x-axis represents the categories inside specific features of the healthcare availability and affordability domain. The categories in each healthcare availability and affordability input feature along with data frequencies are discussed in the appendix in **Table 9**. The y-axis shows the associated SHapley Additive exPlanations values for each category in the same feature of the healthcare availability and affordability domain. The color shades in the color bar represent input categories of different features that interact with the examined feature. Each color shade in the graphic represents a complicated interaction effect between healthcare availability and affordability and other specified attributes including age and heart disease. This provides a clear understanding of the intricate interplay of factors associated with the healthcare availability and affordability domain that affects the model’s output.

**Figure 9.**
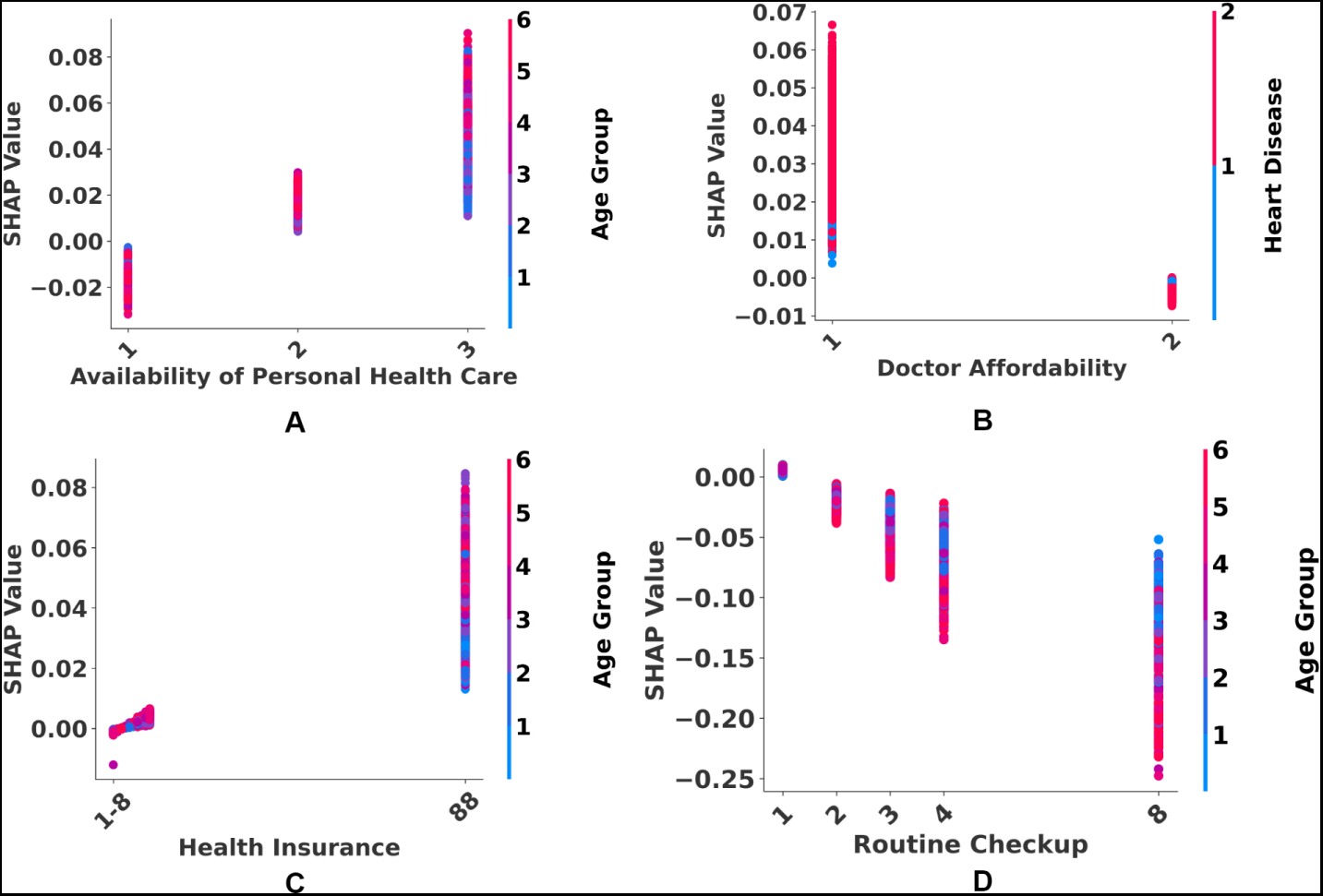
A dependence graphic to represent the predictive effect of each category in healthcare availability and affordability features with interaction effects. SHapley Additive exPlanations (SHAP) values were employed to represent the effect of each category on healthcare availability and affordability features in terms of identifying myocardial infarction (MI) probability including the interaction effects with other features. The x-axis represents the different categories inside a specific feature from healthcare availability and affordability. Besides, the y-axis represents SHAP values for an input category inside that specific feature. The color bar represents different input categories from another feature that interacts with that specific feature.

Identifying health outcomes associated with healthcare availability and affordability by predictive modeling requires careful evaluation of several interacting factors. Each interaction between healthcare availability and affordability and other specified attributes including age and heart disease claimed to have an impact on predicting MI cases if and only if it has positive SHAP values. The **Fig 9** in part **A** shows elderly individuals without access to personal health care have a stronger impact on predictions than younger individuals facing similar constraints. On the other hand, elderly individuals who have access to several different healthcare services have a significant influence on prediction models. Furthermore, the **Fig 9** in part **B** shows individuals experienced financial restraints to see a doctor in need has a significant impact on the prediction. This also interacts with another attribute to show individuals with heart disease in this regard have a minimal prediction impact. Moreover, the **Fig 9** in part **C** shows the effects of elderly individuals without health insurance are greater than those of young individuals in similar situations. Interestingly, the **Fig 9** in part **D** shows elderly individuals, especially those who check within a year have a minimal impact on the prediction.

#### 0.6.5 Analyzing effects and dependencies from socio-demographic domain

The effects and dependencies of each category inside the features from the socio-demographic domain that have an impact on predicting myocardial infarction (MI) cases have been analyzed in this section. This analysis also shows how these selective features from the socio-demographic domain interact with the other features used in this study to influence the prediction of myocardial infarction (MI) probability.

This section shown in **Fig 10** provides a detailed explanation regarding the way features of the socio-demographic factors domain are contributed to predicting MI cases, including the manner in which interact with other features. The x-axis represents the categories inside specific features of the socio-demographic domain. The categories in each socio-demographic input feature along with data frequencies are discussed in the appendix in **Table 10**. The y-axis shows the associated SHapley Additive exPlanations values for each category in the same feature of the socio-demographic domain. The color shades in the color bar represent input categories of different features that interact with the examined feature. Each color shade in the graphic represents a complicated interaction effect between the socio-demographic and other specified attributes including general health, age, and stroke history. This provides a clear understanding of the intricate interplay of factors associated with the healthcare availability and affordability domain that affects the model’s output.

**Figure 10.**
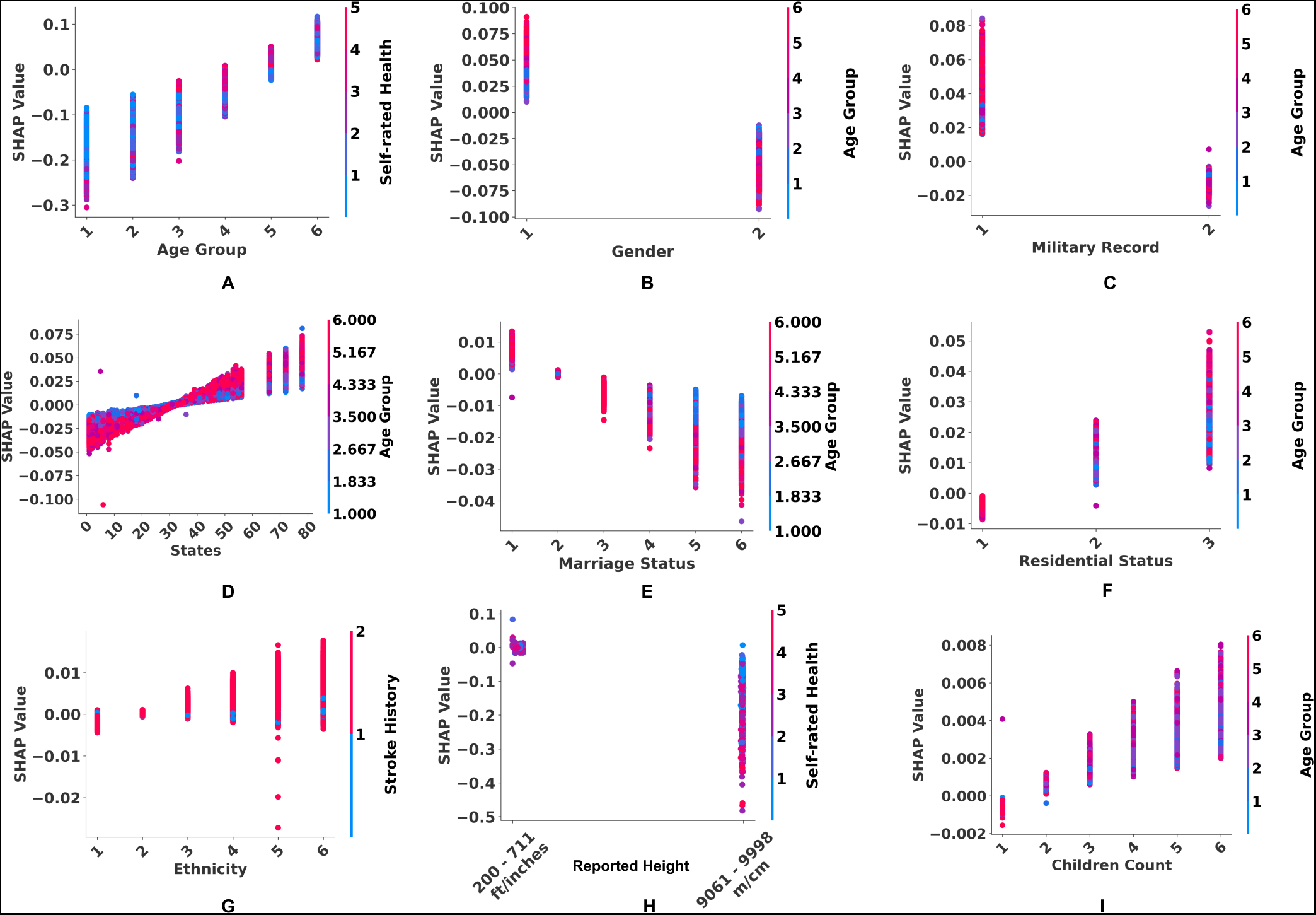
A dependence graphic to represent the predictive effect of each category in socio-demographic features with interaction effects. SHapley Additive exPlanations (SHAP) values were employed to represent the effect of each category in socio-demographic features in terms of identifying myocardial infarction (MI) probability including the interaction effects with other features. The x-axis represents the different categories inside a specific feature from socio-demographics. Besides, the y-axis represents SHAP values for an input category inside that specific feature. The color bar represents different input categories from another feature that interacts with that specific feature.

Identifying health outcomes associated with socio-demographics by predictive modeling requires careful evaluation of several interacting factors. Each interaction between socio-demographic and other specified attributes including general health, age, and stroke history claimed to have an impact on predicting MI cases if and only if it has positive SHAP values. The **Fig 10** in part **A** shows elderly individuals with bad reported health have a stronger contribution to the prediction. Furthermore, the **Fig 10** in part **B** shows elderly males have a significant contribution to the prediction than females. In a similar vein, the **Fig 10** in part **C** shows elderly individuals with a military record have a stronger contribution to the prediction. Accordingly, the **Fig 10** in part **D** shows elderly living all over the states in the US have a significant contribution to the prediction. However, the elderly living in Guam, Puerto Rico, and the Virgin Islands have the strongest contribution to the prediction. The **Fig 10** in part **E** shows marital status is significant with elderly couples having a higher influence on the prediction than those who are younger. Furthermore, the **Fig 10** in part **F** shows living arrangements also impact predicting outcomes, with elderly individuals who lack suitable accommodations and those who live in rented houses having a greater influence than those who are younger. Accordingly, the **Fig 10** in part **G** shows Among certain racial and health groups, minor non-Hispanic, Hispanic American Indian, and Asian individuals have a stronger contribution to the prediction. Additionally, other non-Hispanic races excluding those categorized as White, Black, Asian, and American Indian/Alaskan Native with a stroke history have a minimal contribution to the prediction. Furthermore, the **Fig 10** in part **H** shows individuals reported with negative health have a greater influence than individuals of any height reported with good health. Finally, the **Fig 10** in part **I** shows family size appears as a predictive indicator, with older and middle-aged individuals with more children having a greater influence on prediction than younger individuals with identical family situations.

#### 0.6.6 Analyzing effects and dependencies from personal health habits domain

The effects and dependencies of each category inside the features from the personal health habits domain that have an impact on predicting myocardial infarction (MI) cases have been analyzed in this section. This analysis also shows how these selective features from the personal health habits domain interact with the other features used in this study to influence the prediction of myocardial infarction (MI) probability.

This section shown in **Fig 11** provides a detailed explanation regarding the way features of the personal health habits factors domain are contributed to predicting MI cases, including the manner in which interact with other features. The x-axis represents the categories inside specific features of personal health habits. The categories in each personal health habits input feature along with data frequencies are discussed in the appendix in **Table 11**. The y-axis shows the associated SHapley Additive exPlanations values for each category in the same feature of the personal health habits domain. The color shades in the color bar represent input categories of different features that interact with the examined feature. Each color shade in the graphic represents a complicated interaction effect between personal health habits and other specified attributes including age and general health. This provides a clear understanding of the intricate interplay of factors associated with the personal health habits domain that affects the model’s output.

**Figure 11.**
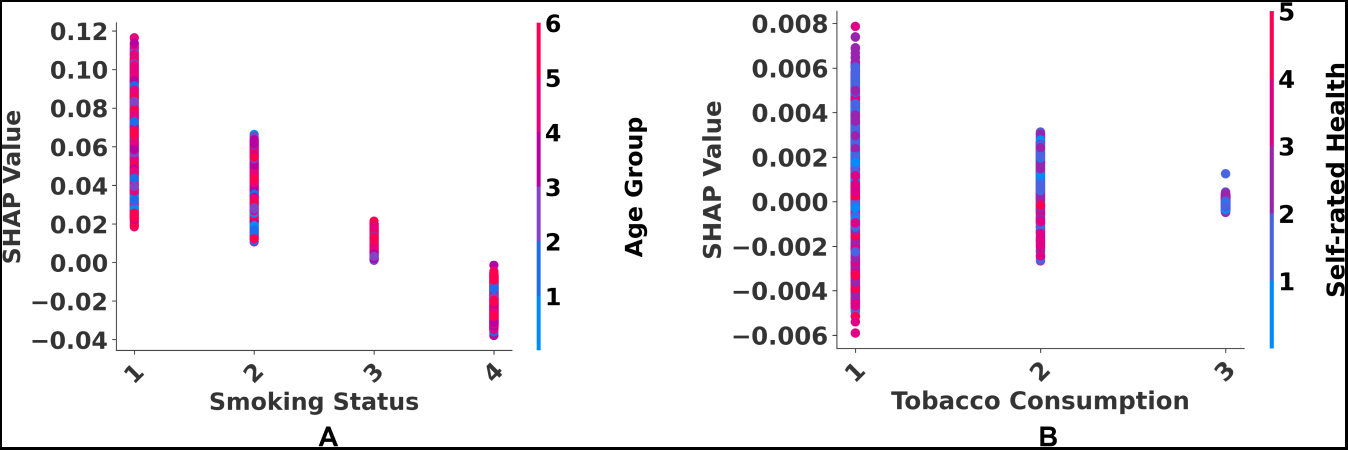
A dependence graphic to represent the predictive effect of each category in personal health habits features with interaction effects. SHapley Additive exPlanations (SHAP) values were employed to represent the effect of each category on personal health habits features in terms of identifying myocardial infarction (MI) probability including the interaction effects with other features. The x-axis represents the different categories inside a specific feature from personal health habits. Besides, the y-axis represents SHAP values for an input category inside that specific feature. The color bar represents different input categories from another feature that interacts with that specific feature.

Identifying health outcomes associated with personal health habits by predictive modeling requires careful evaluation of several interacting factors. Each interaction between personal health habits and other specified attributes including age and general health claimed to have an impact on predicting MI cases if and only if it has positive SHAP values. The **Fig 11** in part **A** shows older individuals have a stronger effect on prediction in three categories: chain-smoking, irregular smoking, and formal smoking compared to younger counterparts. However, the **Fig 11** in part **B** shows When it comes to chewing tobacco products, the dynamics change, with young and middle-aged individuals having a disproportionately larger role in prediction in the two categories of chain chewing and irregular chewing than older individuals.

## Experiment and result analysis

The study conducted in this section includes a thorough analysis of the proposed prediction model using a wide variety of performance measures. This analysis substantiates the model’s critical role in the precise prediction of myocardial infarction (MI) probability from medical survey data by contrasting the model’s results against well-established approaches. Oversampling at various ratios was employed to address the class imbalance. The obtained recall values for each Minority-weighted Sampling ratio were examined using all applied models. Following that, the proposed model’s loss and accuracy at each epoch were assessed to analyze the effect of the class imbalance. Following that, the specificity and sensitivity of all models were evaluated using both raw and balanced data, allowing for a comparison of results. The Area Under the Curve (AUC) for each model was then analyzed in the next section. Finally, correct and incorrect prediction rates were analyzed, offering more insights into the models’ efficacy in regulating class imbalance.

### 0.7 Analyzing recall for different Minority-weighted Sampling ratios

The minority class (MI cases) has experienced a heavy imbalance compared to the majority class (healthy cases). A Minority-weighted Sampling technique has been implemented that oversamples the minority class at different ratios to provide the ideal ratio for each model. Different class distributions have been generated through sampling and recall for each distribution has been analyzed across all the applied models. Specifically, data distribution has been increased gradually from a specific ratio (1:4.69) to a balanced scenario, which provides the changes in the performance of the models across each distribution ratio.

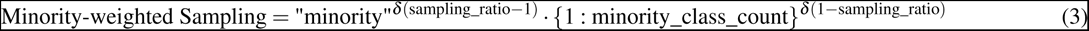

Where: - Minority-weighted Sampling represents the overall strategy used for sampling in the context of dealing with imbalanced datasets. - ‘minority’ indicates that the following sampling strategy is specifically designed for the minority class, which is the class with fewer instances in an imbalanced dataset. - *δ* is a mathematical function that influences the sampling strategy based on the given conditions. It adjusts the impact of the ratio and counts terms in the strategy. - sampling_ratio is a parameter that determines the ratio of sampling. It represents the desired proportion of the minority class instances in the final sampled dataset relative to the majority class instances. - {1 : minority_class_count} specifies the sampling strategy for the minority class. Here, 1 is the label for the minority class, and minority_class_count is the count of samples in the minority class.

The accompanying **Fig 12**. demonstrates the macro-average recall of the class 0 (healthy cases) and class 1 (MI cases). In the trainset, the minority class (MI cases) was oversampled using Minority-weighted Sampling for performing the model training on a balanced data distribution. To identify the oversampling ratio that would provide the best results across the applied approaches, multiple models were trained and evaluated using these specified distributions. The performance of multiple artificial intelligence (AI) models across specific data distributions indicates distinctive trends. Random Under-Sampling (RusBoost), Gaussian Naive Bayes (GNB), and Decision Trees (DT) in particular demonstrated consistent and stable performance throughout the distribution. Adaptive Boosting (AdaBoost), Artificial Neural Networks (ANN), and Random Forests (RF), on the other hand, showed a significant increasing trend, indicating a continuing increase in efficacy over the specified distributions. In contrast, Multilayer Perceptron (MLP) and Convolutional Neural Networks (CNN) produced inconsistent results, implying that performance was subject to fluctuations during the observed distribution. It was identified from the analysis that at the equal training distribution, the proposed ANN model obtained the highest recall among all the applied approaches.

**Figure 12.**
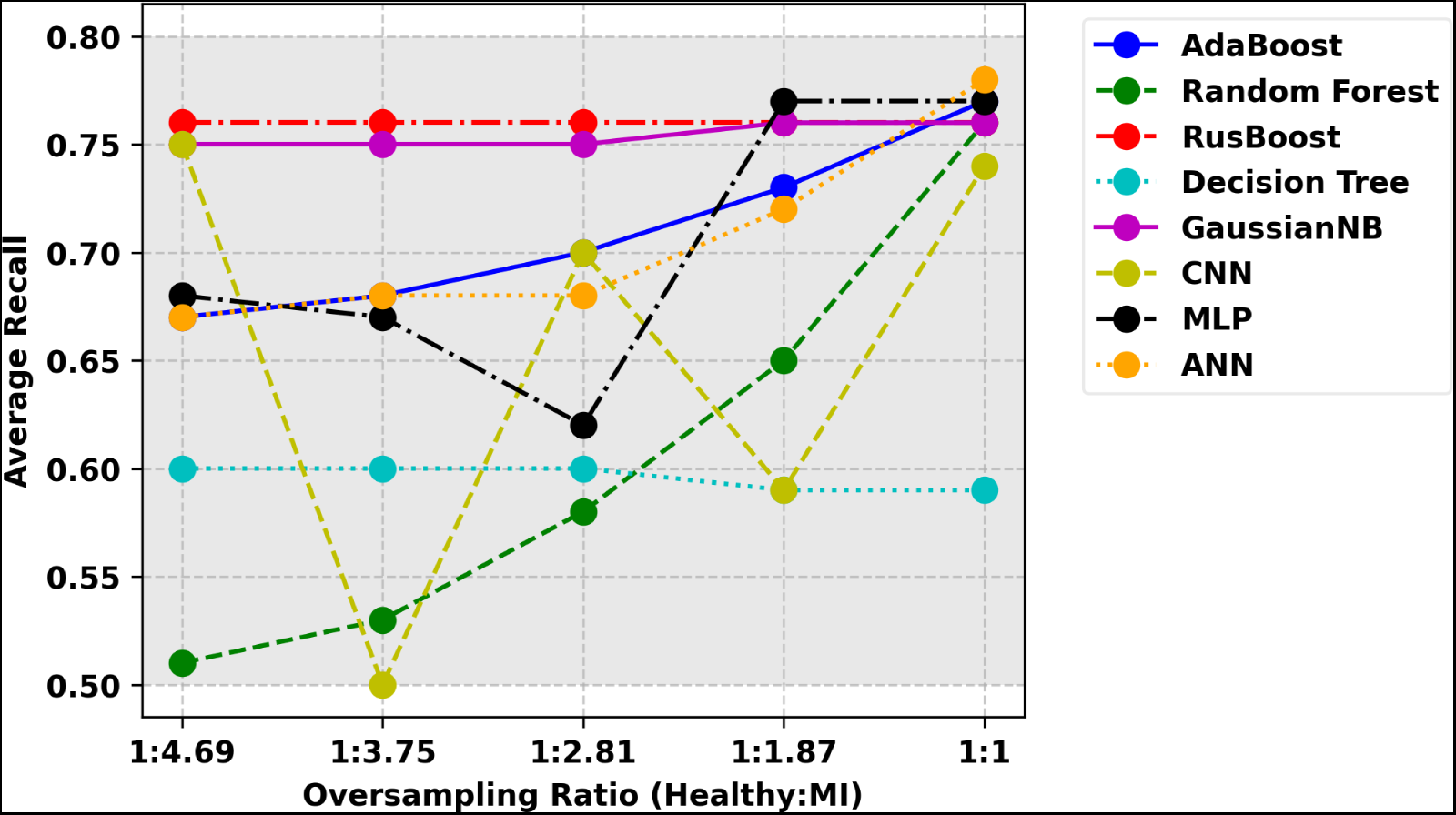
Average recall for all the applied models across each oversampled trainset using Minority-weighted Sampling. Minority class (MI cases) in the train data were oversampled at different ratios using Minority-weighted Sampling and multiple approaches were employed to measure the optimal model for achieving the best outcomes across the given distributions.

### 0.8 Analyzing loss and accuracy of proposed approach

A lightweight Artificial Neural Network (ANN) has been proposed to predict myocardial infarction (MI) probability. The model has 54,273 parameters (weights and biases) in total, of which 53,377 were trained during the training phase and 896 were non-trainable. The total training phases of the proposed model have been conducted through 20 epochs. Loss and accuracy across each epoch have been analyzed to assess the continuous performance of the proposed model in the training and testing phase after handling the class imbalance issue.

**Fig 13**. depicts the training and validation performance of the proposed model over specified epochs. The loss over epochs is depicted with a solid red line indicating training loss and a dashed blue line representing validation loss. The validation curves seem to fluctuate with spikes. The accuracy over epochs is depicted with solid red and dashed blue lines indicating training and validation accuracy, respectively. The accuracy curves have an overall expanding trend with occasional dips.

**Figure 13.**
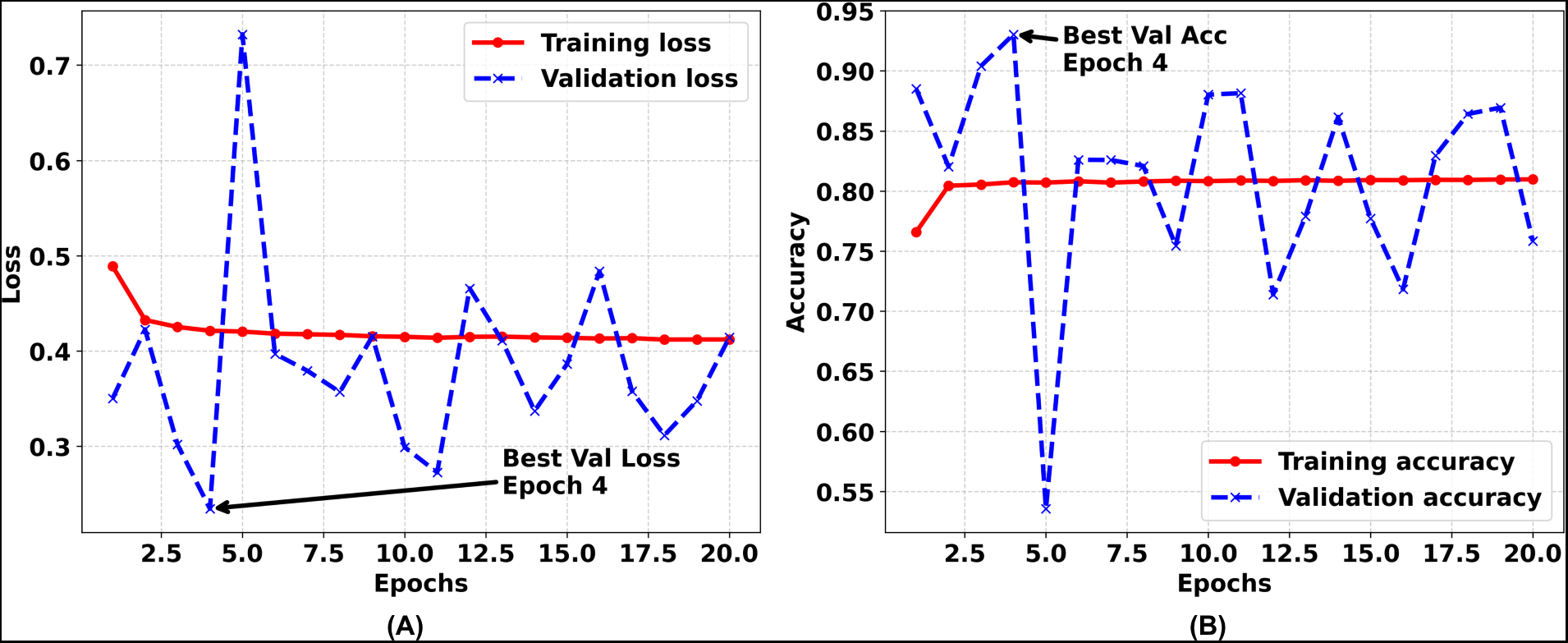
Loss and accuracy of the proposed ANN model with respect to epochs. (A) Visualization of training and validation loss per epoch. (B) Visualization of training and validation accuracy per epoch. The observed changes in the training and validation phases are shown in this section.

### 0.9 Analyzing specificity and sensitivity

Considering the class imbalances, a Minority-weighted Sampling technique has been conducted to improve the performance of the applied models. For each model, performances have been evaluated using both unbalanced data and oversampled balanced data using Minority-weighted Sampling so that improvement through Minority-weighted Sampling can be compared with unbalanced scenarios. To assess the improvement more specifically, specificity and sensitivity have been analyzed to evaluate the capacity of all the proposed models to recognize the true negatives and true positives.

The following formulae were used to compute the specificity and sensitivity.

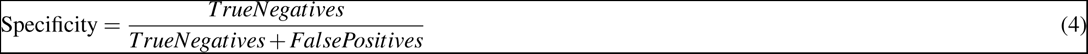

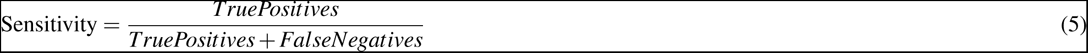

This accompanying **Table 5** presents a comparative analysis of the performance of several artificial intelligence (AI) models in two different training scenarios: training on raw data and balanced training through Minority-weighted Sampling. Specificity, or the capacity to recognize true negatives, and sensitivity, or the capacity to recognize true positives, are the metrics used to evaluate the models^63,64,65,66^. The majority of the models in the training on raw data scenario show high levels of specificity (between 0.96 and 0.98), but these also typically have lower sensitivity values, indicating that these models perform better at accurately categorizing non-events but have trouble identifying actual events. When trained on balanced data, sensitivity values improve for most models. It is noteworthy that, out of all the suggested strategies, the proposed Artificial Neural Network (ANN) model produced the most optimum outcome using the balanced trainset.

**Table 5.**
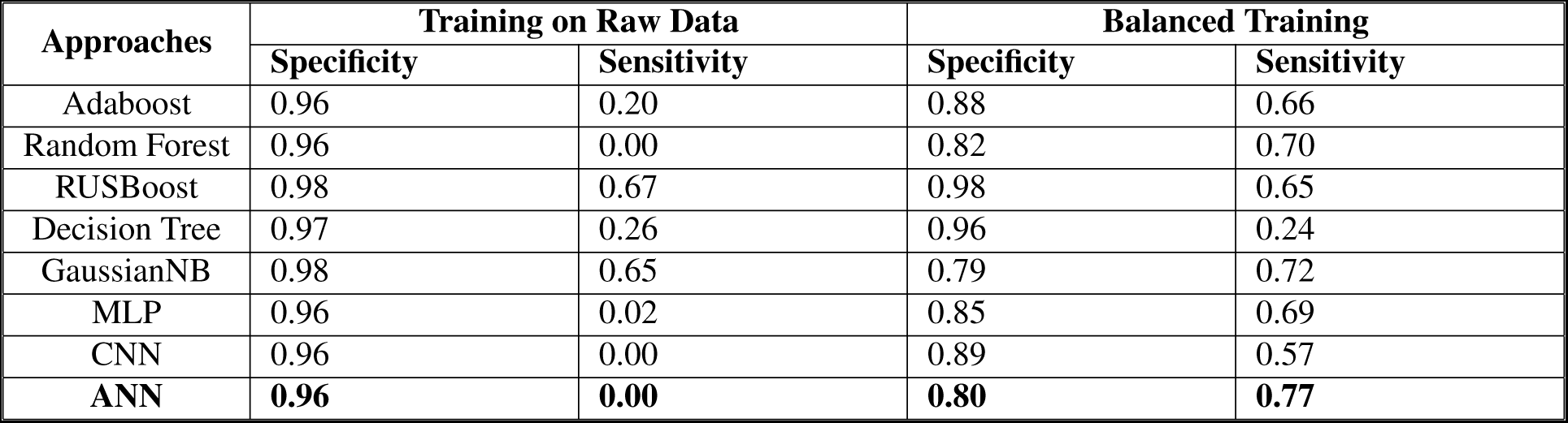
Specificity and sensitivity using both raw data and balanced data across different approaches. Different models were trained on the raw data and accordingly on the oversampled balanced data using Minority-weighted Sampling, and obtained outcomes from both approaches were compared in this section.

| Approaches | Training on Raw Data |  | Balanced Training |  |
| --- | --- | --- | --- | --- |
|  | Specificity | Sensitivity | Specificity | Sensitivity |
| Adaboost | 0.96 | 0.20 | 0.88 | 0.66 |
| Random Forest | 0.96 | 0.00 | 0.82 | 0.70 |
| RUSBoost | 0.98 | 0.67 | 0.98 | 0.65 |
| Decision Tree | 0.97 | 0.26 | 0.96 | 0.24 |
| GaussianNB | 0.98 | 0.65 | 0.79 | 0.72 |
| MLP | 0.96 | 0.02 | 0.85 | 0.69 |
| CNN | 0.96 | 0.00 | 0.89 | 0.57 |
| ANN | **0.96** | **0.00** | **0.80** | **0.77** |

### 0.10 Analyzing Area Under the Curve (AUC)

There are two target classes including healthy cases and MI cases in this study. The Area Under the Curve (AUC) looks at how well the applied models separate these two classes. Specifically, the true positive rates against the false positive rates have been analyzed at different decision thresholds. This analysis has been conducted to assess how many MI cases were correctly identified and how many healthy cases were mistakenly identified as MI cases. The bigger the area, the better-specified model is at distinguishing between the two classes.

The accompanying **Fig 14** depicts the obtained true positive and false positive rates for all the applied models. The AUC evaluations for the various classification models provide details on the performance in terms of sensitivity (true positive rate) and specificity (true negative rate). A higher AUC value indicates a better balance of sensitivity and specificity, indicating a model’s capacity to identify true positives while minimizing false positives. The proposed Artificial Neural Network (ANN) excels in this context, with the highest AUC of 0.87 among all the applied approaches, showing excellent sensitivity and specificity. When compared to the proposed Artificial Neural Network (ANN) model’s performance, the other models possessed relatively poorer performance. Decision Tree (DT), Random Forest (RF), Random Undersampling Boosting (RusBoost), and Gaussian Naive Bayes (GNB) models, all performed rather poorly with AUC scores ranging from 0.60 to 0.76. Comparably, the Convolutional Neural Network (CNN), Adaptive Boosting (AdaBoost), and Multilayer Perceptron (MLP) models performed worse than the suggested ANN model, lagging behind with AUC scores ranging from 0.77 to 0.83. These AUC values provide useful information regarding the way each model handles the sensitivity-specificity trade-off, with higher AUC values indicating more robust performance in both aspects of accurate classification.

**Figure 14.**
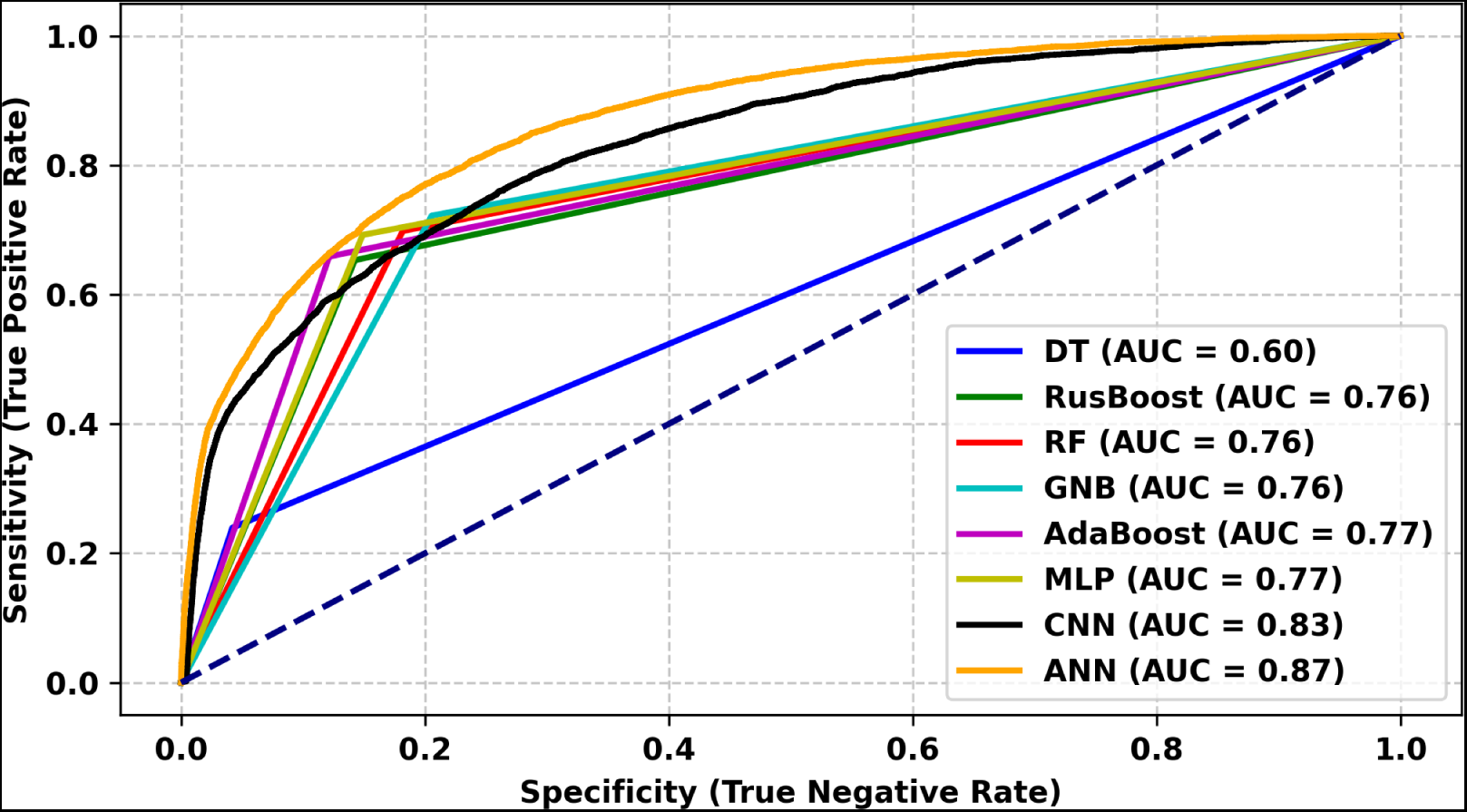
Area Under the Receiver Operating Characteristic (ROC) curve for all the applied approaches. AUC curves for all the applied models are shown in this section to demonstrate the true positive and false positive rates.

### 0.11 Analyzing correct and incorrect prediction rates

There are two instances in this study including negative (healthy cases) and positive (MI cases). The confusion matrices have been analyzed in this study to provide the relative distribution of predictions by showing the proportions of true positive, true negative, false positive, and false negative predictions. This normalization has been conducted to assess the model’s performance in a more contextually meaningful way, especially to deal with imbalanced datasets. In imbalanced datasets, a normalized confusion matrix helps by showing how well a model works for each class, considering its unequal sizes, making it easier to understand and improve performance, especially for the minority class.

The accompanying **Fig 15** depicts the confusion matrix to represent the correct and incorrect prediction rate. The findings of the study indicated a concerning trend in the performance of the several models used, with high specificity but noticeably poor sensitivity a crucial parameter when it comes to determining individuals at risk of having a myocardial infarction (MI). Such models are expected to be highly effective at identifying possible myocardial infarction (MI) instances, however, most of the examined models were not very effective at it. Despite its complexity, deep neural networks were inefficient for this study as increasing the network’s depth increased the issue of prioritizing the majority class above balanced predictions. However, a lightweight Artificial Neural Network model outperformed the performance by producing more promising outcomes for both healthy and MI cases.

**Figure 15.**
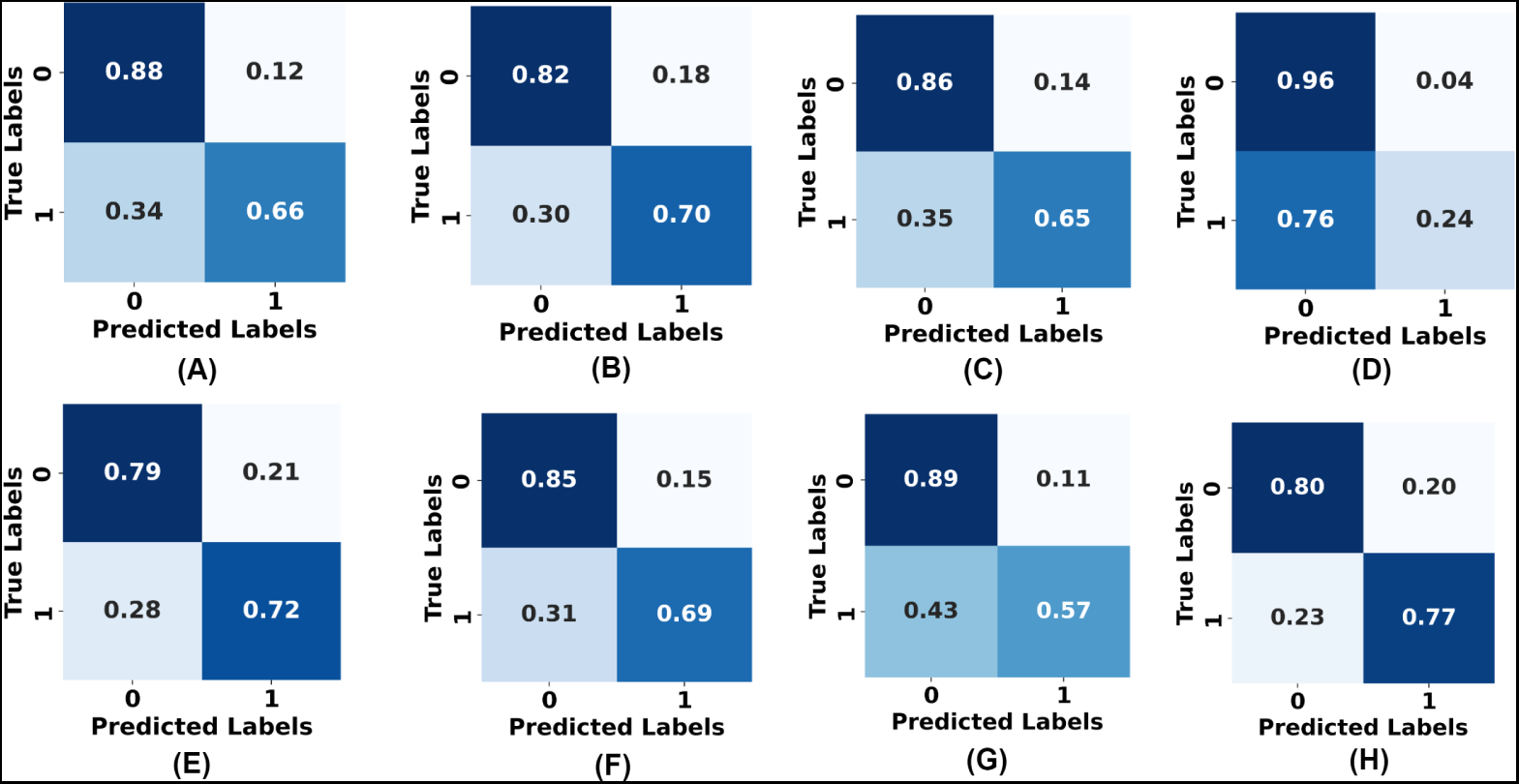
Confusion matrix for all the applied models. (A) Confusion matrix of the Adaptive Boosting model. (B) Confusion matrix of the Random Forest model. (C) Confusion matrix of the Random Under-Sampling model. (D) Confusion matrix of the Decision Tree model. (E) Confusion matrix of the Gaussian Naive Bayes model. (F) Confusion matrix of the Multilayer Perceptron model. (G) Confusion matrix of the Convolutional Neural Network model. (H) Confusion matrix of the proposed Artificial Neural Network model. The representation of correct and incorrect prediction rates for both prediction classes.

## Discussion

In conclusion, this study has highlighted a number of significant gaps in the literature on the use of artificial intelligence in cardiovascular diagnostics, particularly in the context of cardiovascular disease prediction^26,27,28,32^. The proposed approach has fluctuating performance in predicting myocardial infarction (MI) due to the class imbalance issue and the proposed model’s sensitivity to it. For artificial intelligence (AI) models, class imbalance can have a major impact on validation loss and accuracy. When there is a significant difference in the number of samples across classes, the model could be biased towards the majority class, resulting in decreased accuracy for the minority class^67^. In terms of validation loss, the model might have difficulty accurately classifying instances from the minority class, resulting in increased loss owing to frequent classifications. Due to the observed imbalance in the class distribution within the target column which included a disproportionately high number of healthy cases most applied models exhibited a bias in favor of this majority class. As the model struggles to generalize, this results in a fluctuating validation loss curve with spikes. The Minority-weighted Sampling was employed to resolve the class imbalance issue, as seen by better validation loss and accuracy in subsequent epochs of the proposed model, overcoming the early challenges imposed by the class imbalance. This indicates that balancing the dataset improves the models’ ability to provide a generalized outcome. Prior studies^28,29,30,51,52,53,54,55^ claimed excellent accuracy but lacked in-depth examination, raising questions about the usefulness of their models. Simply having high accuracy does not guarantee strong specificity and sensitivity^26,27^. For example, a prior^26^ showed high accuracy (0.89) but obtained sensitivity was low (0.27). Accordingly, another prior study^27^ showed high accuracy (0.9157) but similarly obtained low specificity (0.5261). This approach relies heavily on sensitivity (recall for MI cases), particularly in predicting myocardial infarction (MI). On the other hand, prior study^48^ showed high specificity (0.909) and sensitivity (0.926), although they tested on limited and balanced datasets (healthy cases: 33 records, heart disease cases: 41 records). Real-world scenarios contain uneven data and prior study^26,27^ has shown that substantial imbalances can cause AI models to perform badly. For example, a study conducted by Mamun et al.^27^ using a large dataset (319795 records) obtained unbalanced specificity (0.5261) and sensitivity (0.9232). Accordingly, an artificial intelligence model was proposed by Akkaya et al.^26^ that tested on a large and unbalanced dataset (healthy: 51884 records, heart disease: 4175 records), but the obtained unbalanced specificity (0.94) and sensitivity (0.27). In this study, we explicitly tested our model on imbalanced scenarios with a large number of samples (healthy cases: 107829 records and MI cases: 4964 records) and were able to obtain balanced specificity (0.80) and sensitivity (0.77) in realistic conditions.

By employing explainable AI (XAI) in the proposed model, it was possible to identify significant risk indicators for myocardial infarction (MI). Individuals who have previous angina or coronary heart disease history were observed to have the most significant impact on predicting myocardial infarction (MI) cases. Angina or coronary heart disease in the past indicates that the arteries may still be obstructed which could increase the likelihood of myocardial infarction (MI). Hence, the obtained comparative probability for heart disease history above 0.4 indicates that it could be a major risk factor for myocardial infarction (MI). Accordingly, individuals who have a previous stroke history were observed to have a significant impact on predicting myocardial infarction (MI) cases. A history of stroke could raise the possibility of vascular abnormalities, which could increase the risk of heart disease. Hence, the obtained comparative probability for stroke history above 0.25 indicates previous stroke history could be an important risk factor for myocardial infarction (MI). Further, diabetes in the elderly seemed to have a significant impact on predicting MI cases. High levels of sugar in the blood over time could make the blood vessels less flexible and more prone to blockages of blood. Diabetes could be another important risk factor for the cardiovascular health of the elderly. Accordingly, bronchitis seemed to have a significant impact on myocardial infarction (MI) prediction. The inflammation and infection in the airways could lead to increased strain on the heart. Hence, bronchitis could be another important risk factor for myocardial infarction (MI). Individuals who are underweight seemed to have a stronger impact on the prediction of myocardial infarction (MI) probability. Park et al.^31^ conducted research that also indicated underweight is a significant risk factor for cardiovascular disease. Surprisingly, in the elderly group, those who are underweight seemed to be more impactful in predicting myocardial infarction (MI) probability. The aging process could cause elements like plaque to build up in the arteries, rendering elderly individuals more susceptible to heart attacks. Hence, the predictive influence of underweight elderly makes the low body mass index (BMI) an important factor in predicting myocardial infarction (MI). Smokers have a greater influence on myocardial infarction (MI) probability prediction. Heavy smoking could damage blood vessels, reducing oxygen supply to the heart and contributing to the formation of blood clots. Hence, smoking could be an important risk factor for myocardial infarction (MI). However, formal smokers (comparative probability: 0.02) seemed to have less comparative probability of myocardial infarction (MI) than chain smokers (comparative probability: 0.10). Quitting smoking could improve blood vessel function and decrease the formation of blood clots. Hence, it indicates quitting smoking could reduce the risk of myocardial infarction (MI).

The proposed Artificial Neural Network (ANN) model shows good interpretability measured by Shapley values, indicating a higher incidence of myocardial infarction (MI) among individuals dealing with chronic medical conditions like high blood pressure, high cholesterol, diabetes, bronchitis, asthma, kidney diseases, low body mass index (BMI), and a history of previous heart disease or stroke. Additionally, the model highlights a strong relationship between myocardial infarction (MI) and socio-demographic variables, economic standing, healthcare accessibility, individual health practices, and disability statuses, such as blindness, errand difficulty, and mobility limitations. This thorough research adds to the comprehension of the complex interplay between the many risk factors for myocardial infarction (MI) and its epidemiology, opening up new possibilities for focused intervention and preventive measures. Additionally, the thorough approach used in this study offers a solid framework for advanced research in the area of medical AI to ensure substantial specificity and sensitivity. The careful selection of relevant variables, thorough data preprocessing, and development of architecture have demonstrated the potential for developing accurate and resilient AI models, even in the context of challenging data issues. The use of explainable AI approaches and sensitivity analysis improves the model’s transparency and trustworthiness, making it more useful for healthcare applications.

## Data Availability

https://www.cdc.gov/brfss/index.html

## A Appendix

**Table 6.**
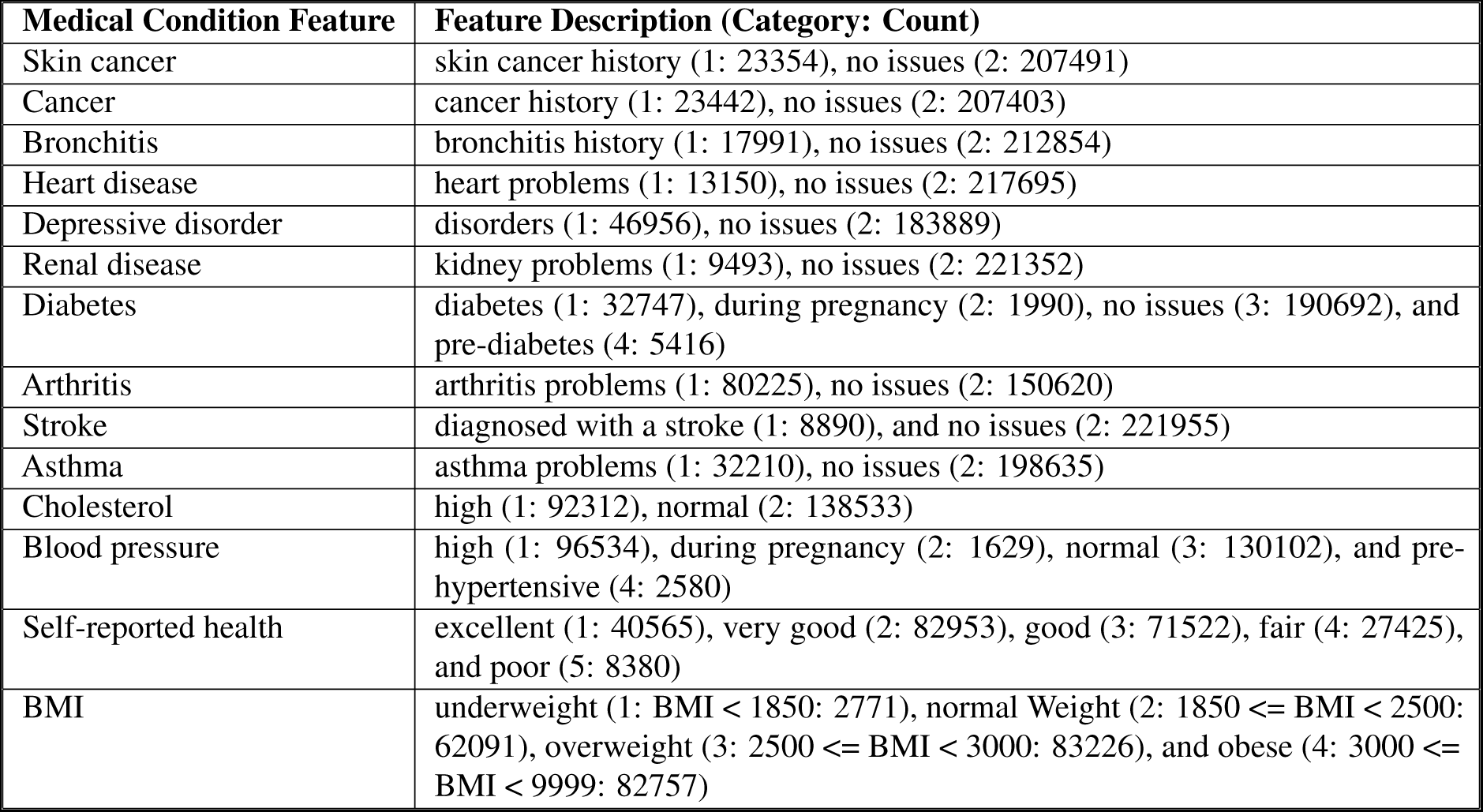
Features from the medical conditions input domain. The categories and values of each medical condition feature were described in this section.

**Table 7.** Features from the impairment status input domain. The categories and values of each impairment status feature were described in this section.

| **Impairment Status Feature** | **Feature Description (Category: Count)** |
| --- | --- |
| Difficulty seeing | difficulties (1: 10630), no issues (2: 220215) |
| Difficulty dressing or bathing | difficulties (1: 8471), no issues (2: 222374) |
| Difficulty walking or climbing stairs | difficulties (1: 35476), no issues (2: 195369) |
| Difficulty doing errands alone | physical, mental, or emotional condition that causes difficulties (1: 14859), no issues (2: 215986) |

**Table 8.**
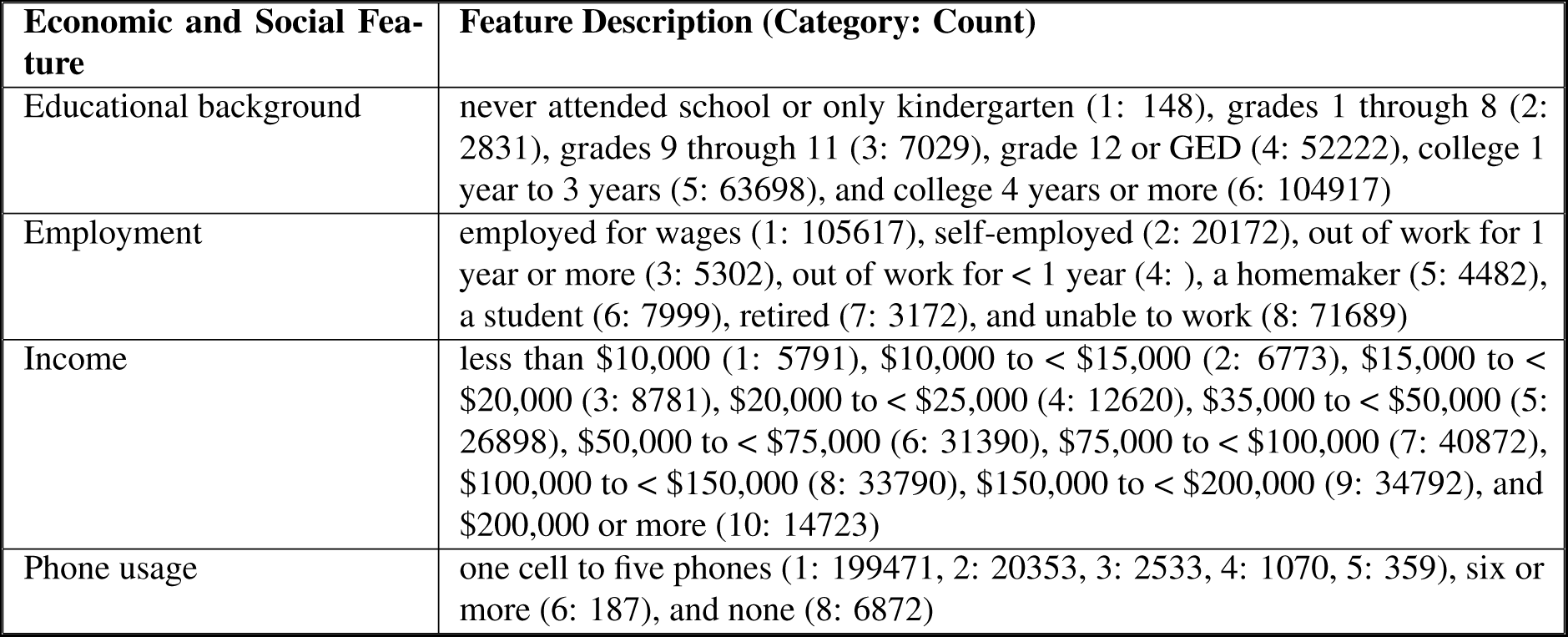
Features from the economic and social status input domain. The categories and values of each economic and social feature were described in this section.

**Table 9.**
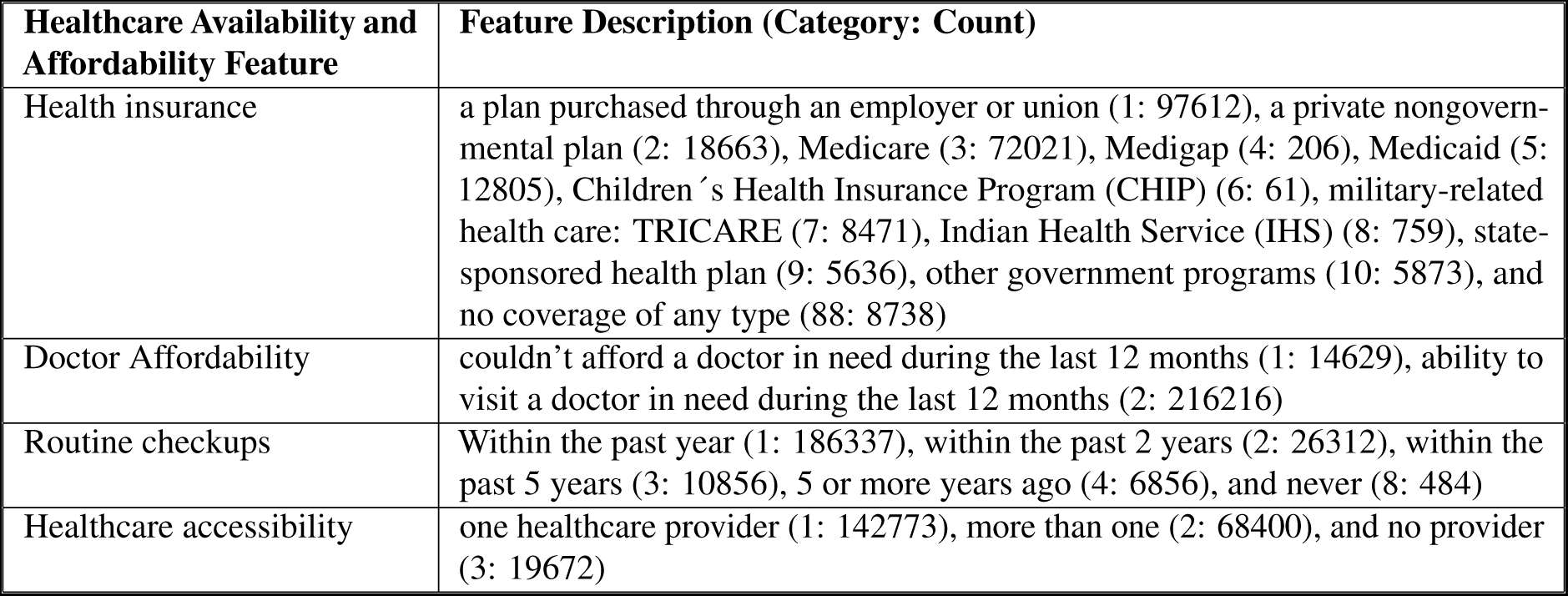
Features from the healthcare availability and affordability input domain. The categories and values of each healthcare availability and affordability status feature were described in this section.

| **Healthcare Availability and Affordability Feature** | **Feature Description (Category: Count)** |
| --- | --- |
| Health insurance | a plan purchased through an employer or union (1: 97612), a private nongovernmental plan (2: 18663), Medicare (3: 72021), Medigap (4: 206), Medicaid (5: 12805), Children's Health Insurance Program (CHIP) (6: 61), military-related health care: TRICARE (7: 8471), Indian Health Service (IHS) (8: 759), state-sponsored health plan (9: 5636), other government programs (10: 5873), and no coverage of any type (88: 8738) |
| Doctor Affordability | couldn't afford a doctor in need during the last 12 months (1: 14629), ability to visit a doctor in need during the last 12 months (2: 216216) |
| Routine checkups | Within the past year (1: 186337), within the past 2 years (2: 26312), within the past 5 years (3: 10856), 5 or more years ago (4: 6856), and never (8: 484) |
| Healthcare accessibility | one healthcare provider (1: 142773), more than one (2: 68400), and no provider (3: 19672) |

**Table 10.** Features from the socio-demographic input domain. The categories and values of each socio-demographic feature were described in this section.

| Socio-demographic Feature | Feature Description (Category: Count) |
| --- | --- |
| Marital status | married (1: 130370), divorced (2: 31447), widowed (3: 23436), separated (4: 4203), never married (5: 32815), and a member of an unmarried couple (6: 8574) |
| Residential status | own a home (1: 175557), rent a home (2: 48390), and other arrangements (3: 6898) |
| Military record | military background (1: 30580), no records in this regard (2: 200265) |
| Self-reported height | 200 - 711 ft/inches: 193529, 9061 - 9998 meters/centimeters: 37316 |
| Ethnicity | White, Non-Hispanic (1: 178345), Black, Non-Hispanic (2: 17034), Asian, Non-Hispanic (3: 5654), American Indian/Alaskan Native, Non-Hispanic (4: 3657), Hispanic (5: 18627), and Other race, Non-Hispanic (6: 7528) |
| States | Alabama (1: 2530), Alaska (2: 2886), Arizona (4: 5998), Arkansas (5: 2701), California (6: 4195), Colorado (8: 5530), Connecticut (9: 4274), Delaware (10: 2012), District of Columbia (11: 1699), Georgia (13: 4031), Hawaii (15: 4865), Idaho (16: 3377), Illinois (17: 1540), Indiana (18: 4893), Iowa (19: 5138), Kansas (20: 9558), Kentucky (21: 2701), Louisiana (22: 2693), Maine (23: 4505), Maryland (24: 8537), Massachusetts (25: 3578), Michigan (26: 4844), Minnesota (27: 8864), Mississippi (28: 2387), Missouri (29: 6330), Montana (30: 3576), Nebraska (31: 8979), Nevada (32: 1477), New Hampshire (33: 3472), New Jersey (34), New Mexico (35), New York (36), North Carolina (37), North Dakota (38), Ohio (39), Oklahoma (40), Oregon (41), Pennsylvania (42), Rhode Island (44), South Carolina (45), South Dakota (46), Tennessee (47), Texas (48), Utah (49), Vermont (50), Virginia (51), Washington (53), West Virginia (54), Wisconsin (55), Wyoming (56), Guam (66), Puerto Rico (72), Virgin Islands (78), New Jersey (34: 3716), New Mexico (35: 3706), New York (36: 18588), North Carolina (37: 2698), North Dakota (38: 3190), Ohio (39: 7653), Oklahoma (40: 2489), Oregon (41: 2434), Pennsylvania (42: 3276), Rhode Island (44: 3033), South Carolina (45: 4953), South Dakota (46: 3737), Tennessee (47: 2546), Texas (48: 5441), Utah (49: 5497), Vermont (50: 3542), Virginia (51: 5170), Washington (53: 7367), West Virginia (54: 3976), Wisconsin (55: 3945), Wyoming (56: 2408), Guam (66: 1004), Puerto Rico (72: 2562), and Virgin Islands (78: 744) |
| Spoken language | English (1: 224597), Spanish (2: 6248) |
| Gender | male (1: 110171), female (2: 120674) |
| Children count | no children (1: 170652), one child (2: 25089), two children (3: 21733), three children (4: 8836), four children (5: 3065), and five or more children (6: 1470) |
| Age | youth (1: aged 18 to 24: 6576), early adults (2: aged 25 to 34: 20819), middle-aged adults (3: aged 35 to 44: 32238), mature adults (4: aged 45 to 54: 38643), senior adults (5: aged 55 to 64: 49428), and elderly (6: aged 65 or over: 83141) |

**Table 11.** Features from the personal health habits input domain. The categories and values of each personal health habit status feature were described in this section.

| **Personal Health Habits Feature** | **Feature Description (Category: Count)** |
| --- | --- |
| Alcohol consumption | no drinking habits (1: 216629), no habits in this regard (2: 14216) |
| Tobacco consumption | regular consumer (1: 20721), occasional consumer (2: 7810), former smoker (3: 66401), and never (4: 135913) |
| Smoking status | heavy smoker (1: smoke every day: 20721), light smoker (2: smoke some days: 7810), quit smoking (3: 66401), and never smoked (4: 135913) |
| Exercise | participating in sports and exercises (1: 179310), no habits in this regard (2: 51535) |
| Fruit consumption | 101 - 199 days: 118918, 201 - 299 weeks: 72322, 300 less than a month: 1529, 301 - 399 month/year: 32564, and 555 never: 5512 |

**Table 12.** Features from the preventive health service input domain. The categories and values of each preventive health service feature were described in this section.

| **Preventive Health Services Feature** | **Feature Description (Category: Count)** |
| --- | --- |
| HIV test | tested for HIV (1: 81396), never tested for HIV (2: 149449) |
| Vaccination | flu vaccine during the past 12 months (1: 128096), didn't take any flu vaccine during the past 12 months (2: 102749) |

## Notes

### Competing Interest Statement

The authors have declared no competing interest.

### Funding Statement

This study did not receive any funding

### Author Declarations

Behavioral Risk Factor Surveillance System https://www.cdc.gov/brfss/index.html

